# Inferring community transmission of SARS-CoV-2 in the United Kingdom using the ONS COVID-19 Infection Survey

**DOI:** 10.1101/2023.10.24.23297454

**Authors:** Ruth McCabe, Gabriel Danelian, Jasmina Panovska-Griffiths, Christl A. Donnelly

**Affiliations:** Department of Statistics, University of Oxford; National Institute for Health and Care Research Health Protection Research Unit in Emerging and Zoonotic Infections; United Kingdom Health Security Agency; The Queen’s College, University of Oxford; The Pandemic Sciences Institute, University of Oxford; MRC Centre for Global Infectious Disease Analysis, Imperial College London

**Keywords:** Effective reproduction number, instantaneous growth rate, SARS-CoV-2, COVID-19, United Kingdom, ONS COVID-19 Infection Survey, surveillance

## Abstract

Key epidemiological parameters, including the effective reproduction number, *R*(*t*), and the instantaneous growth rate, *r*(*t*), generated from an ensemble of models, have been informing public health policy throughout the COVID-19 pandemic in the four nations of the United Kingdom of Great Britain and Northern Ireland (UK). However, estimation of these quantities became challenging with the scaling down of surveillance systems as part of the transition from the “emergency” to “endemic” phase of the pandemic.

The Office for National Statistics (ONS) COVID-19 Infection Survey (CIS) provided an opportunity to continue estimating these parameters in the absence of other data streams. We used a penalised spline model fitted to the ONS CIS test positivity estimates to produce a smoothed estimate of the prevalence of SARS-CoV-2 positivity over time. The resulting fitted curve was used to estimate the “ONS-based” *R*(*t*) and *r*(*t*) across the four nations of the UK. Estimates produced under this model are compared to government-published estimates with particular consideration given to the contribution that this single data stream can offer in the estimation of these parameters.

Depending on the nation and parameter, we found that up to 77% of the variance in the government-published estimates can be explained by the ONS-based estimates, demonstrating the value of this singular data stream to track the epidemic in each of the four nations. We additionally find that the ONS-based estimates uncover epidemic trends earlier than the corresponding government-published estimates.

Our work shows that the ONS CIS can be used to generate the key COVID-19 epidemics across the four UK nations. This is not intended as an alternative to ensemble modelling, rather it is intended as a potential solution to the aforementioned challenge faced by public health officials in the UK in early 2022.

## Introduction

Key epidemiological parameters, including the effective reproduction number, *R*(*t*), and the instantaneous growth rate, *r*(*t*), have been used to inform public health policy throughout the COVID-19 pandemic.^1–4^ Estimation of these quantities by public health officials in the United Kingdom (UK) has relied on an ensemble of models which encompass a range of data sources and assumptions.^5^ These parameters are traditionally estimated using case data as a proxy for the infection incidence curve,^6,7^ but methods have also been developed to estimate these parameters from other sources, such as hospitalisations^8^ and genomic data.^9^ The four nations of the UK (England, Scotland, Wales and Northern Ireland, herein ordered by population size) were recognised globally as having comprehensive SARS-CoV-2 testing surveillance systems,^10–12^ comprising of widescale community testing,^13^ nationwide surveys of infection,^14,15^ genomic data,^16,17^ and wastewater surveillance,^18^ but these were largely scaled down from their peak capacity as part of the transition from an “emergency” to “endemic” state in the first half of 2022.^19–22^ Consequently, estimation of *R*(*t*) and *r*(*t*), particularly using an ensemble model approach, became more challenging due to the reduction in available data streams.

The Office for National Statistics (ONS) COVID-19 Infection Survey (CIS)^14^ was a primary means by which SARS-CoV-2 transmission was tracked within the UK. This COVID-19 testing study invited members of randomly selected private households across the UK to complete polymerase chain reaction (PCR) tests, regardless of symptoms or behaviour. The ONS CIS continued for one year after the cessation of community testing, until its “pause” in March 2023,^23^ and in October 2023 it was announced that a similar study would be undertaken for the upcoming 2023/24 winter period.^24^ Although the survey can estimate incidence of infection, this lags the main metric of interest, the percentage of the population testing positive for SARS-CoV-2 infection, which is used as a proxy for the prevalence of infection in each nation of the UK. These data, widely regarded as “gold-standard” due to the random sampling methods underpinning them, provided an opportunity to continue estimating *R*(*t*) and *r*(*t*) after the scaling down of many of the surveillance systems described above.

By adapting the methods deployed by another community surveillance survey with randomly-selected participants, the REal-time Assessment of Community Transmission (REACT) study,^15,25^ this paper presents a model to estimate *R*(*t*) and *r*(*t*) directly from publicly available ONS CIS test positivity estimates in each nation of the UK. The estimates produced under this model are compared to government-published ensemble estimates, to assess the validity of this method to track the spread of SARS-CoV-2 in the absence of other surveillance data. In particular, we consider the level of contribution that this single data stream can offer in the estimation of these parameters. This methodology is then used to provide estimates of *R*(*t*) and *r*(*t*) in each nation of the UK until the initial pause of the ONS CIS in the first quarter of 2023.

## Methods

### Estimating R(t) and r(t) using the ONS COVID-19 Infection Survey (CIS)

#### The ONS CIS

The ONS CIS was the only long-term SARS-CoV-2 testing study in randomly selected households encompassing all four nations of the UK. In brief, private households were randomly selected across each nation, irrespective of factors such as members displaying symptoms or having contact with a known case, and household members aged over 2 years were invited to complete multiple PCR tests over time. The random sampling produced estimates that were unaffected by test-seeking behaviour and public availability of diagnostic tests. Further details regarding sampling can be found in ^26^.

The primary outcome of interest was the estimated percentage of people testing positive for SARS-CoV-2, derived from the number of positive tests out of the total tests completed and post-stratified by key variables, such as gender and ethnicity. The publication of corresponding estimates of incidence of infection, based on the primary outcome (prevalence of test positivity), was lagged by a couple of weeks, as this is substantially more complex to estimate, but ceased to be published from June 2022.^27^ Initially, the CIS had overlapping reporting windows of approximately 10 – 14 days but reporting settled into approximately weekly windows.

Publicly-available data are provided as point estimates with 95% credible intervals, derived from the post-stratification model.^26^ The survey commenced on different dates in each nation (Table 1) and there is some heterogeneity in the reporting windows across nations, but was “paused” on 13 March 2023 in England, Scotland and Wales and on 7 March 2023 in Northern Ireland (with the last publication (at time of writing) on 24 March 2023).^28^ In all settings, the midpoint of the reporting window is taken as the “date” of the observation, for the purposes of model fitting.

**Table 1:** Availability of estimates used in this study: test positivity from the ONS CIS survey and official estimates of the effective reproduction number and growth as published by the UK government. The study period for each nation is determined as the beginning date of the ONS CIS until the initial “pause” date of the ONS CIS (March 2023), while the period of overlap states the period for which both ONS-based estimates and government-published estimates are available for comparison.

| Nation | ONS CIS coverage | Parameter | Government-published estimate coverage | Period of overlap |
| --- | --- | --- | --- | --- |
| England | 27/04/20 – 13/03/23 | Reproduction number | 29/05/20 – 23/12/22 | 29/05/20 – 23/12/22 |
|  |  | Growth rate | 12/06/20 – 23/12/22 | 12/06/20 – 23/12/22 |
| Scotland | 03/10/20 – 13/03/23 | Reproduction number | 28/05/20 – 08/12/22 | 03/10/20 – 08/12/22 |
|  |  | Growth rate | 18/06/20 – 08/12/22 | 03/10/20 – 08/12/22 |
| Wales | 27/07/20 – 13/03/23 | Reproduction number | 28/05/20 – 09/12/22 | 27/07/20 – 09/12/22 |
|  |  | Growth rate | 12/06/20 – 27/12/21 | 27/07/20 – 27/12/21 |
| Northern Ireland | 11/09/20 – 07/03/23 | Reproduction number | 26/05/20 – 31/05/22 | 11/09/20 – 31/05/22 |
|  |  | Growth rate | 09/06/20 – 17/11/20 | 11/09/20 – 17/11/20 |

In October 2023, the UKHSA and the ONS announced a “Winter COVID-19 study” (WCIS) running from November 2023 – March 2024, with similar aims to the ONS CIS of ascertaining prevalence of SARS-CoV-2 infection in communities across the UK.^24^ However, as these data are not available at the time of writing (October 2023) and their exact format is currently unknown, our study only covers the first, continuous period of the ONS CIS from 2020 to the end of March 2023. Nonetheless, the implications of our methodology for the newly announced WCIS are outlined in the Discussion.

#### Modelling R(t) and r(t)

A two-step approach is used to estimate *R*(*t*) and *r*(*t*) from the ONS CIS, with these estimates denoted by 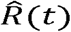 and 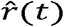, respectively, and referred to as “ONS-based” estimates. The methods are primarily adapted from those presented in Eales et al.^25^ and akin to those in ^9,29,30^. Specifically, a penalised spline model is fit to the ONS CIS test positivity estimates to produce a smoothed estimate of the prevalence of SARS-CoV-2 positivity over time before the epidemiological parameters of interest are estimated directly from the resulting curve. Each step is described in turn below.

#### Step 1: fitting a spline to the ONS CIS test positivity data

The model used by the REACT study^25^ was taken as the basis for spline-fitting in this study and is explained here briefly. The REACT-1 data consist of the number of positive (*Y*_*t*_) and total (*N*_*t*_) tests on each day *t* of the study period, allowing test positivity of SARS-CoV-2 to be naively estimated as 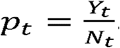. Smoothed test positivity, defined as 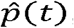, is estimated via a linear combination of B-splines up to the 2^nd^ degree, defined by a sequence of equidistant knots distributed throughout the period under consideration. Specifically:

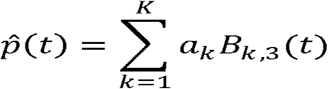

where *B*_*k*,3_(*t*) represents the *k*th 3rd order B-spline (see Supplementary Material) and *a*_*k*_ are the model coefficients, defined by a second-order random-walk prior distribution. The binomial likelihood is then parameterised by the observed number of total tests, *N*_*t*_, and the The estimated test positivity, 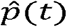, at each time point.

Although the principal idea is the same, several adjustments were required to adapt the REACT spline model to ONS test positivity estimates. First, an additional sampling step was required to capture the uncertainty in the ONS estimates, which are provided in a different format to the REACT-1 data. Let *μ*_*t*_ denote the central ONS estimate of SARS-CoV-2 test positivity at time *t*, with *l*t and *u*t denoting the corresponding lower and upper limits of the credible interval, respectively. The variance of the estimate can be estimated as 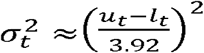. For each time *t, π*_*t*_, the test positivity, is considered a random variable following a Beta distribution parameterised using *μ*_*t*_ and 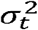 as shown:

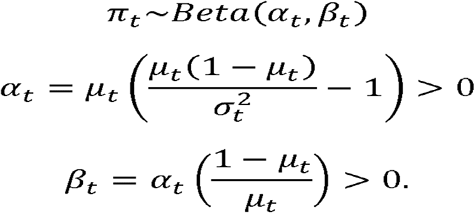

The derivation of the expressions for *α*_*t*_ and *β*_*t*_ is shown in the Supplementary Material. For every iteration, *i*, our model begins by sampling 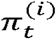 for every time point *t* ahead of the construction of the B-splines and transforming this value onto the unconstrained logit scale:

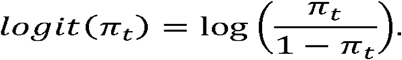

Second, the weekly timescale of the ONS CIS data, compared to the daily timescale of the REACT-1 study data, also necessitated adjustments to the model. In this instance, a first-order random walk was deemed a more appropriate prior distribution for the model coefficients due to the rapidly changing epidemic dynamics in these weekly data. Furthermore, knots could not be placed at the REACT-1-optimised value of every five days. Rather, a sensitivity analysis of the placing of knots was undertaken by considering scenarios in which the number of equidistant knots was equal to a percentage of the total data points, specifically 20%, 30% and 40%, balancing capturing sufficient information but without overfitting.

The final adaptation was to use a Normal likelihood function, again due to the different format of the data, based on the results of a simulation study, with details set out in the Supplementary Material. Bringing everything together:

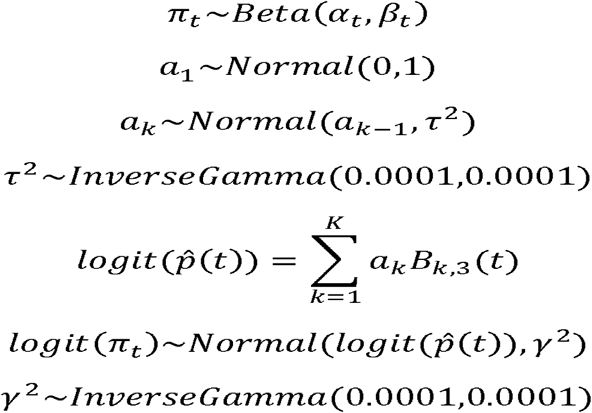

This model was fitted using a No-U-Turn sampler^31^, implemented in STAN, with 4 chains, each with 20,000 iterations *R*(*t*) and *r*(*t*), a burn-in of 2,000 iterations. To ensure smooth estimates of *R*(*t*) and *r*(*t*), the coefficients from the model are used to estimate 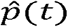 at a granular time step (hundredths of one day).

#### Step 2a: Estimating R(t)

The effective reproduction number, *R*(*t*), is estimated using the renewal equation^6,32^:

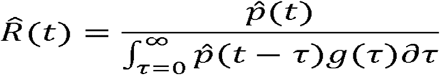

where *g*(.) denotes the distribution of the generation time, defined as the time between the infections of infector-infectee pairs. The integral is approximated by summation.

The generation time distribution has been shown to change over time with the emergence of new variants.^33^ As such, estimates of *g*(.) used in this study were extracted from the literature for four distinct time periods defined by epidemic waves: Wildtype, Alpha, Delta and Omicron. Each study distribution was parameterised using data from the UK and thus is assumed to be applicable to this setting. The transition dates between time periods were determined by the earliest date at which 50% of the daily tests were attributable to the emerging variant according to Our World in Data.^34^ The distributions are summarised in Table 2.

**Table 2:**
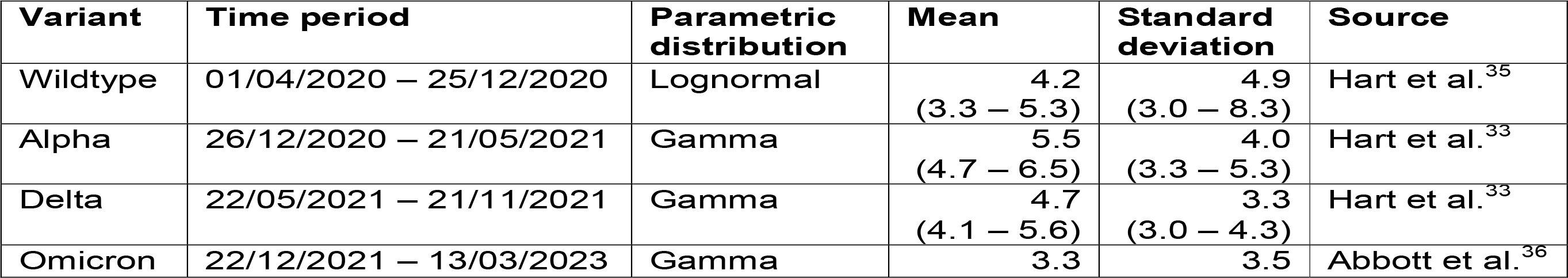
Parameterisations of the generation time distribution used in the analysis. Variant time periods are defined by the earliest date at which 50% of the daily tests were attributable to the emerging variant according to Our World in Data.^34^ Uncertainty was not characterised for the estimates obtained for the Omicron wave.

#### Step 2b: Estimating r(t)

The instantaneous growth rate, *r*(*t*), is approximated as follows:

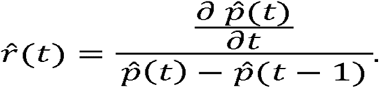

The quantities of interest, 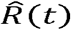 and 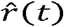, are estimated for all posterior realisations of the spline model, with results presented as the median and the 2.5% and 97.5% quantiles throughout.

### Analysis of 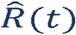 and 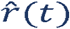

#### Government-published estimates

Estimates of *R*(*t*) and *r*(*t*) for SARS-CoV-2 in each of the four nations were produced for and published by the government from early in the pandemic for either weekly or biweekly time periods.^5,13^ The estimates were derived from an ensemble of (up to 14) independently run models as part of a cross-government and academic modelling hub that comprised the UK Health Security Agency (UKHSA) Epidemiological Ensemble team and Scientific Pandemic Influenza Group on Modelling, Operational sub-group (SPI-M-O).^5,37^ These models encompassed a range of different assumptions and data streams as set out in ^5^. Among the data streams used, officially reported cases and hospital admissions were the most common, but there were also two models which fit to the ONS CIS data.^38–40^ Consequently, there is a degree of inbuilt dependency between the government-published estimates and the estimates obtained in this study, as demonstrated below.

Ensemble model outputs were combined into a single estimate with associated uncertainty, using an established method derived by the Defence Science and Technology Laboratory (DSTL).^41^ These estimates are publicly available for download and are presented as 90% confidence intervals for the period, without any central estimate.^13^ We apply the weekly or biweekly estimates to each day in the corresponding time period, and approximate a central estimate by taking the mid-point of the upper and lower bounds, herein denoted by 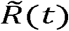 and 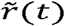 for the effective reproduction number and instantaneous growth rate, respectively. These estimates are herein referred to as “government-published” estimates.

There is heterogeneity in the availability of the government-published estimates, depending on the parameter and nation under consideration (Table 1). For example, official estimates of the growth rate in Northern Ireland are only available from June to November 2020. All estimates for each nation ceased to be published by 23 December 2022.^42^

#### Comparison of ONS-based and government-published estimates

The ONS-based estimates 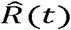 and 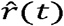 are initially presented from the beginning of the survey period in each nation until the end of 2022, as this is when the government-published estimates ceased to be publicly available for both parameters and for all four nations (Table 1). The periods for which the ONS-based estimates can be compared to government-published estimates for each parameter and nation combination are set out in Table 1. Due to the short period of government published estimates for the growth rate in Northern Ireland, ONS-based estimates of this parameter cannot be compared in any meaningful way to the corresponding official estimates and are thus omitted from our analysis.

We use three metrics to consider the relationship between the (median) ONS-based [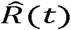 and 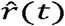] and government-published [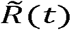 and 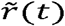] estimates of the parameters of interest, each of which are now presented. These are herein referred to as the “comparison metrics”.

First, we use linear regression models of the form:

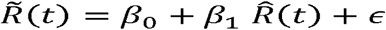

and

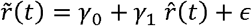

to assess the level of variation within the government-published estimates explained by the ONS-based estimates. This is captured by:

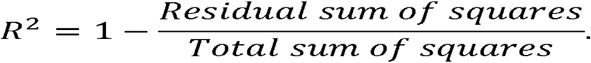

Second, we consider the Spearman rank correlation, selected due to the lack of assumptions regarding the distributions of the data.

Finally, the parameters of interest in this paper are sometimes interpreted dichotomously by policymakers, for example, to assess whether the epidemic is either growing or shrinking. As such, the proportion of point estimates over the study period for which the modelled and official estimates are on the same side of the “growth thresholds”, 0 for the growth rate and 1 for the effective reproduction number, over time is also presented as the third and final metric for comparison. This quantity is herein referred to as the “agreement proportion”.

A sensitivity analysis was used to examine the potential of a time-lag between estimates, assessed via the three comparison metrics. Specifically, lags between plus and minus 20 days from the default values were considered. A positive lag of *L* days indicates that the ONS-based estimates of day *D*, 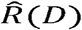 and 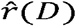, are compared with the government-published estimates at day *D* +*L*, 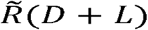 and 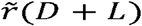, and vice versa. In the main plots, we present our ONS-based estimates alongside the government-published estimates for the time lag and knot value under which *R*^2^ is maximised. A comparison of estimates without any time lags are presented in the Supplementary Material.

#### 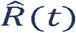 and 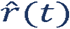 in the first quarter of 2023

Finally, 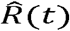 and 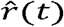 are produced for the first quarter of 2023 by fitting the spline model to ONS CIS estimates from 1 November 2022 until the initial “pause” of the survey on 13 March 2023 in England, Scotland and Wales and on 7 March 2023 in Northern Ireland. These are presented for information but without comparison, due to the lack of availability of the government-published estimates in this period.

All data and code used are available from: https://github.com/ruthmccabe/ons-test-positivity-model.

## Results

Figure 1 to Figure 4 present the results for England, Scotland, Wales and Northern Ireland, respectively. Across the board, we can demonstrate strong agreement between our ONS-based estimates and the government-published estimates for both parameters. Depending on nation and parameter, up to 77% of the variance in the government-published estimates can be explained by the ONS-based estimates, providing evidence of the suitability of this singular data stream to track the epidemic in each of the four nations (Table 3). Similarly, we observed high maximum values of the Spearman rank correlation, ranging between 0.72 and 0.87 (Table 3).

**Table 3:** Maximum values of the three comparison metrics used to assess the relationship between the ONS-based and government-published estimates of R(t) and r(t) for each nation. The value of the metric is provided alongside the lag* (in days) and the percentage of knots** out of the total data points for which this maximum value is produced. For any instance in which there were multiple combinations producing the same maximum value, the lag closest to 0 and lowest percentage of knots was selected for presentation here.

|  |  | Effective reproduction number |  |  | Instantaneous growth rate |  |  |
| --- | --- | --- | --- | --- | --- | --- | --- |
| | | $R^2$ | Spearman correlation | Agreement proportion | $R^2$ | Spearman correlation | Agreement proportion |
| England | Value | 0.71 | 0.85 | 0.81 | 0.77 | 0.87 | 0.82 |
|  | Lag* | -8 | -6 | -10 | -9 | -7 | -7 |
|  | Knots** | 30% | 30% | 40% | 30% | 30% | 40% |
| Scotland | Value | 0.74 | 0.86 | 0.79 | 0.69 | 0.83 | 0.80 |
|  | Lag* | -8 | -8 | -4 | -11 | -10 | -7 |
|  | Knots** | 40% | 40% | 40% | 40% | 40% | 40% |
| Wales | Value | 0.66 | 0.81 | 0.79 | 0.67 | 0.82 | 0.42 |
|  | Lag* | -12 | -10 | -10 | -15 | -14 | -13 |
|  | Knots** | 40% | 40% | 40% | 40% | 40% | 40% |
| Northern Ireland | Value | 0.52 | 0.72 | 0.52 |  |  |  |
|  | Lag* | 9 | 9 | 10 |  |  |  |
|  | Knots** | 40% | 40% | 40% |  |  |  |

**Figure 1:**
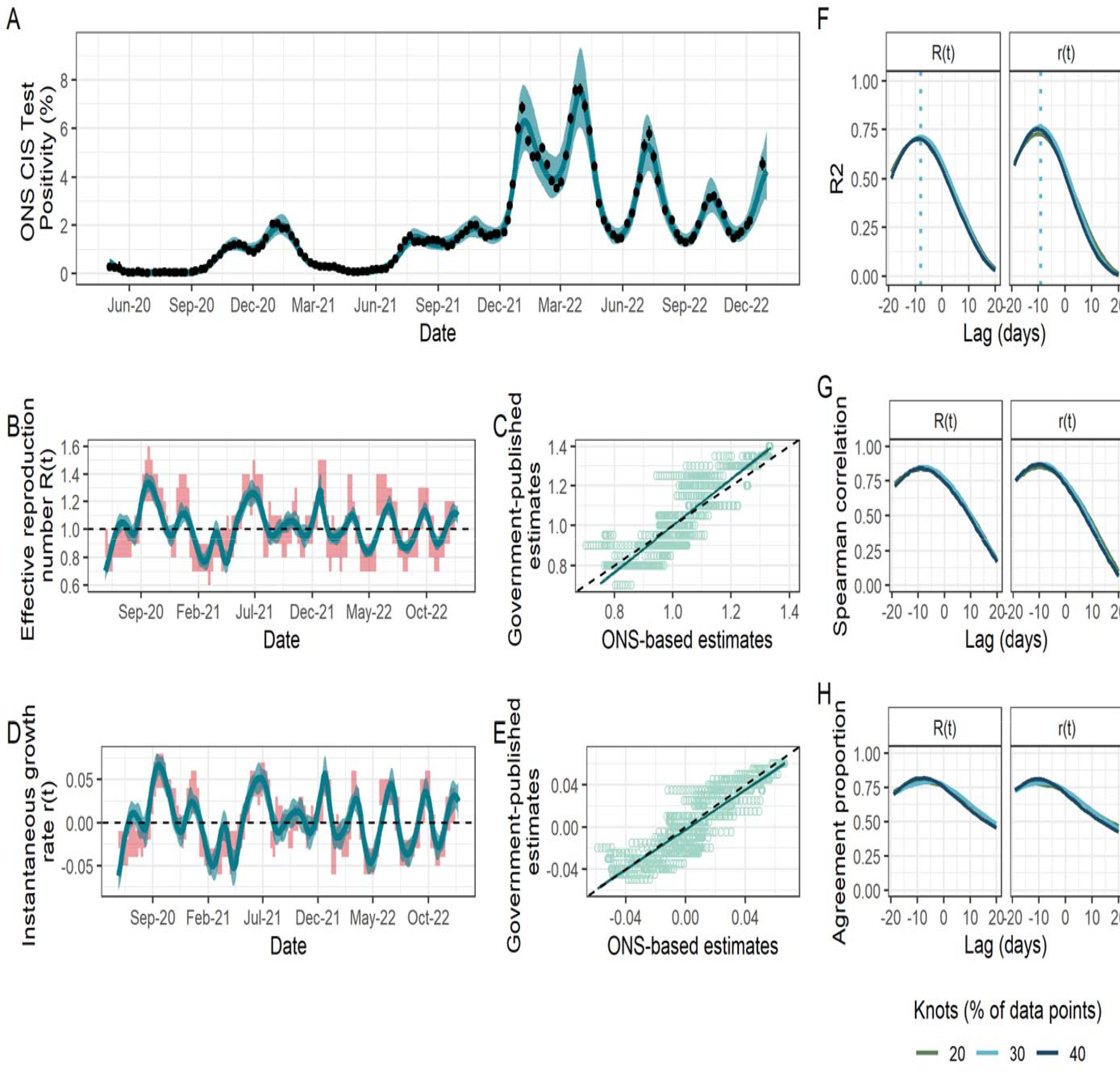
Fit to ONS CIS data, resulting estimates of R(t) and r(t), and comparison metrics for England. (A) Spline model fit 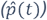 (blue; median line with 95% credible intervals) to data (black points; ONS point estimate with 95% credible intervals) with the number of knots totalling 30% of the total data points. (B) The ONS-based estimated effective reproduction number 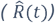 (blue; median line with 95% credible intervals) alongside the government-published estimates 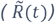 (red; 90% confidence intervals), lagged by -8 days. The black dashed line highlights the epidemic growth threshold of 1. (C) Scatterplot (light green points) showing the relationship between the ONS-based and government-reported estimates of R(t) and r(t), with the latter lagged by -8 days. The dark green line shows the trend line from the linear model (with dark green shading corresponding to 95% confidence intervals) regressing the lagged government-published estimates on the ONS-based estimates. Rows of points are due to the limited precision available (due to rounding before publication) for government-published estimates. (D) The ONS-based estimated instantaneous growth rate 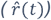 (blue; median line with 95% credible intervals) alongside the government-published estimates 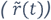 (red; 90% confidence intervals), lagged by -9 days. The dashed line highlights the epidemic growth threshold of 0. (E) Scatterplot (light green points) showing the relationship between the ONS-based and government-reported estimates of r(t), with the latter lagged by -9 days. The dark green line shows the trend line from the linear model (with dark green shading corresponding to 95% confidence intervals) regressing the lagged government-published estimates on the ONS-based estimates. Rows of points are due to the limited precision available (due to rounding before publication) for government-published estimates. (F) – (H) Results of the three metrics used to assess the agreement between ONS-based and government-published estimates under different lags and for models fitted with a different number of knots, taken as a percentage of the total data points. (F) R^2^. Dotted lines indicate the lag for which this metric is maximised for each parameter and is thus what is applied to the government-based estimates in panels (B) – (E). (G) Spearman rank correlation. (H) Agreement proportion.

**Figure 2:**
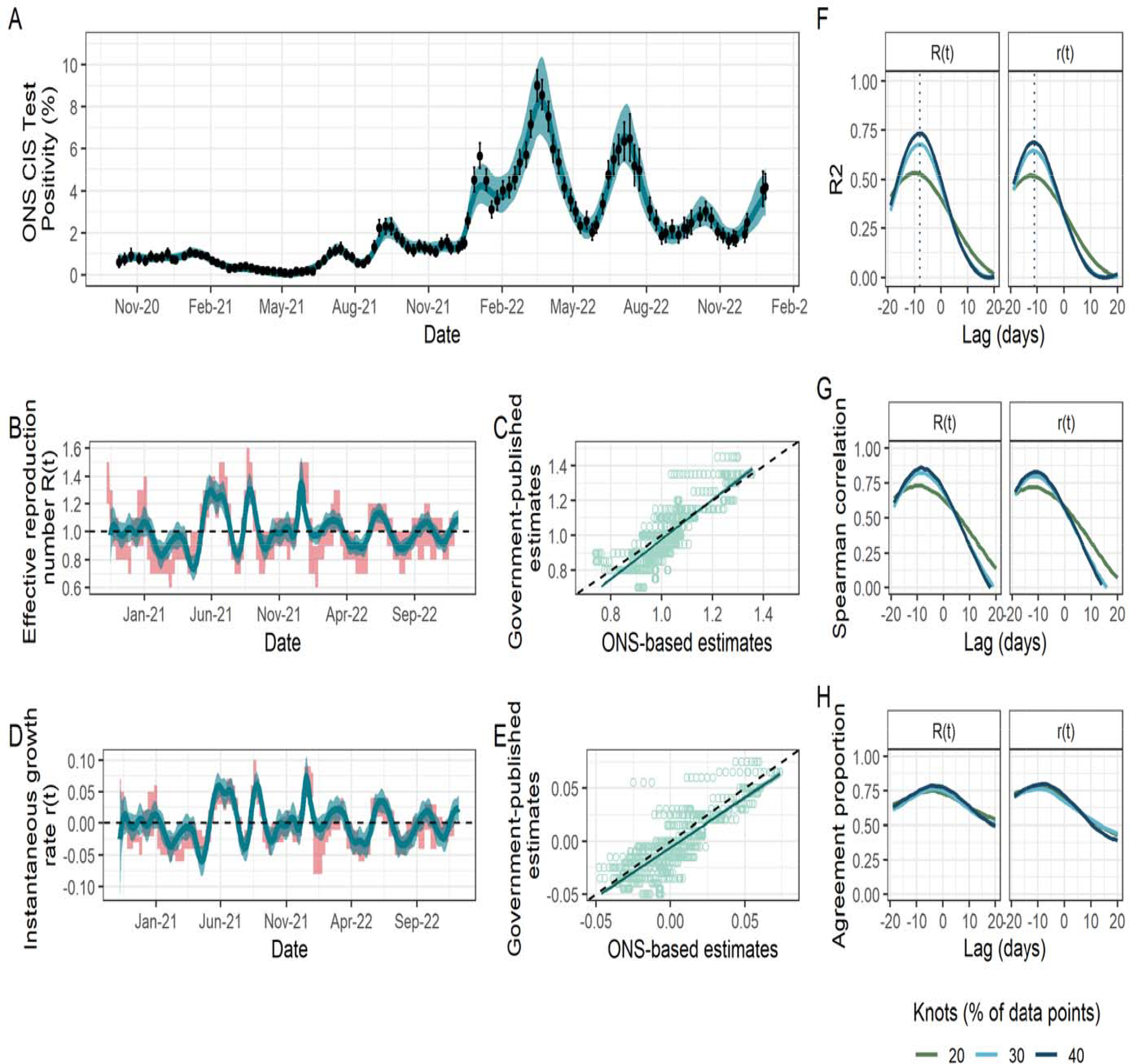
Fit to ONS CIS data, resulting estimates of R(t) and r(t), and comparison metrics for Scotland. (A) Spline model fit 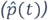 (blue; median line with 95% credible intervals) to data (black points; ONS point estimate with 95% credible intervals) with the number of knots totalling 40% of the total data points. (B) The ONS-based estimated effective reproduction number 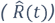 (blue; median line with 95% credible intervals) alongside the government-published estimates 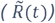 (red; 90% confidence intervals), lagged by -8 days. The black dashed line highlights the epidemic growth threshold of 1. (C) Scatterplot (light green points) showing the relationship between the ONS-based and government-reported estimates of R(t), with the latter lagged by -8 days. The dark green line shows the trend line from the linear model (with dark green shading corresponding to 95% confidence intervals) regressing the lagged government-published estimates on the ONS-based estimates. Rows of points are due the limited precision available (due to rounding before publication) for government-published estimates. (D) The ONS-based estimated instantaneous growth rate 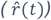 (blue; median line with 95% credible intervals) alongside the government-published estimates 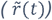 (red; 90% confidence intervals), lagged by -11 days. The dashed line highlights the epidemic growth threshold of 0. (E) Scatterplot (light green points) showing the relationship between the ONS-based and government-reported estimates of r(t), with the latter lagged by -11 days. The dark green line shows the trend line from the linear model (with dark green shading corresponding to 95% confidence intervals) regressing the lagged government-published estimates on the ONS-based estimates. Rows of points are due to the limited precision available (due to rounding before publication) for government-published estimates. (F) – (H) Results of the three metrics used to assess the agreement between ONS-based and government-published estimates under different lags and for models fitted with a different number of knots, taken as a percentage of the total data points. (F) R^2^. Dotted lines indicate the lag for which this metric is maximised for each parameter and is thus what is applied to the government-based estimates in panels (B) – (E). (G) Spearman rank correlation. (H) Agreement proportion.

**Figure 3:**
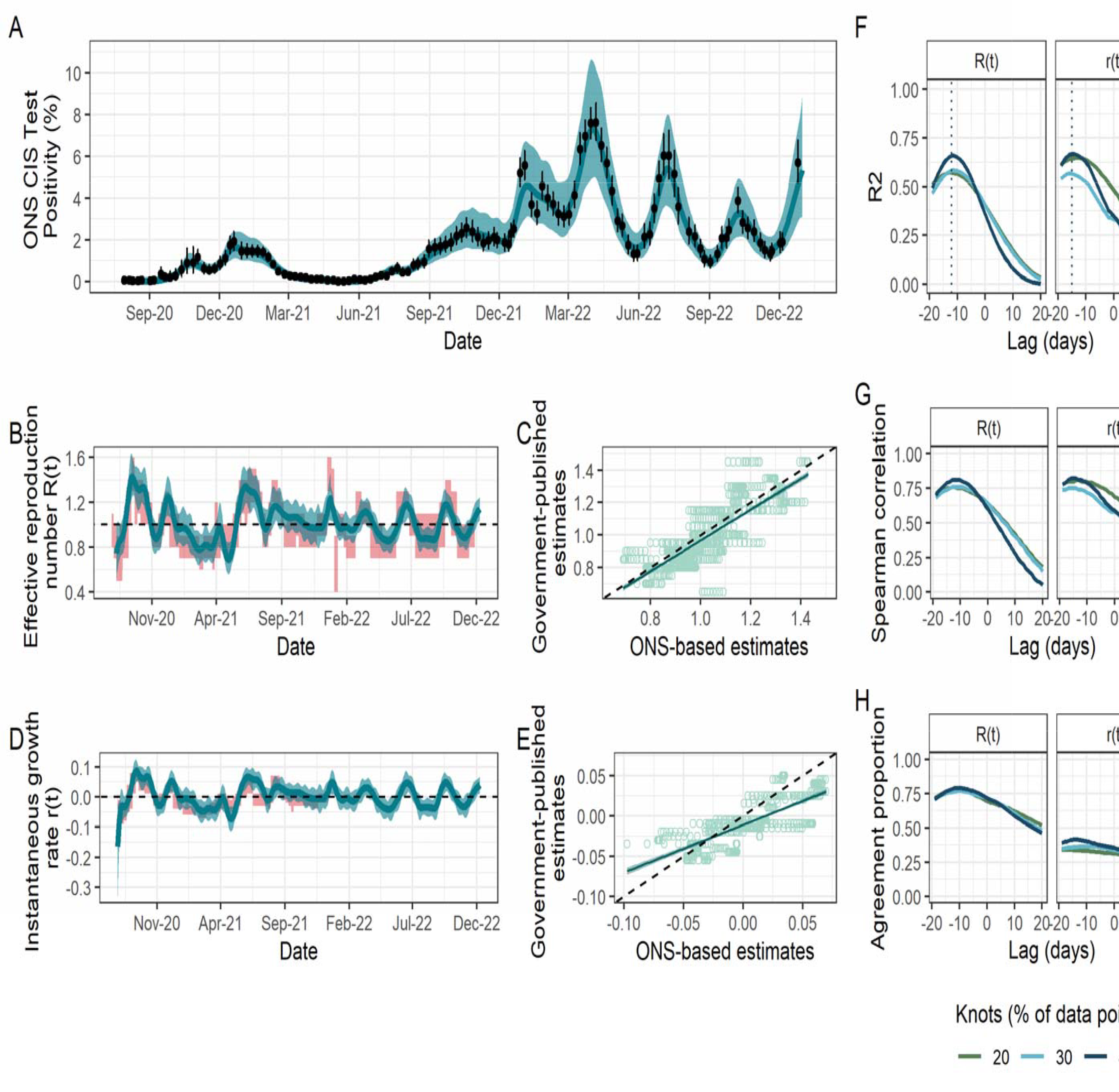
Fit to ONS CIS data, resulting estimates of R(t) and r(t), and comparison metrics for Wales. (A) Spline model fit 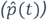 (blue; median line with 95% credible intervals) to data (black points; ONS point estimate with 95% credible intervals) with the number of knots totalling 40% of the total data points. (B) The ONS-based estimated effective reproduction number 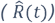 (blue; median line with 95% credible intervals) alongside the government-published estimates 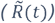 (red; 90% confidence intervals), lagged by - 12 days. The black dashed line highlights the epidemic growth threshold of 1. (C) Scatterplot (light green points) showing the relationship between the ONS-based and government-reported estimates of R(t), with the latter lagged by -12 days. The dark green line shows the trend line from the linear model (with dark green shading corresponding to 95% confidence intervals) regressing the lagged government-published estimates on the ONS-based estimates. Rows of points are due to the limited precision available (due to rounding before publication) for government-published estimates. (D) The ONS-based estimated instantaneous growth rate 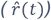 (blue; median line with 95% credible intervals) alongside the government-published estimates 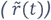 (red; 90% confidence intervals), lagged by -15 days. The dashed line highlights the epidemic growth threshold of 0. (E) Scatterplot (light green points) showing the relationship between the ONS-based and government-reported estimates of r(t), with the latter lagged by -15 days. The dark green line shows the trend line from the linear model (with dark green shading corresponding to 95% confidence intervals) regressing the lagged government-published estimates on the ONS-based estimates. Rows of points are due to the limited precision available (due to rounding before publication) for government-published estimates. (F) – (H) Results of the three metrics used to assess the agreement between ONS-based and government-published estimates under different lags and for models fitted with a different number of knots, taken as a percentage of the total data points. (F) R^2^. Dotted lines indicate the lag for which this metric is maximised for each parameter and is thus what is applied to the government-based estimates in panels (B) – (E). (G) Spearman rank correlation. (H) Agreement proportion.

**Figure 4:**
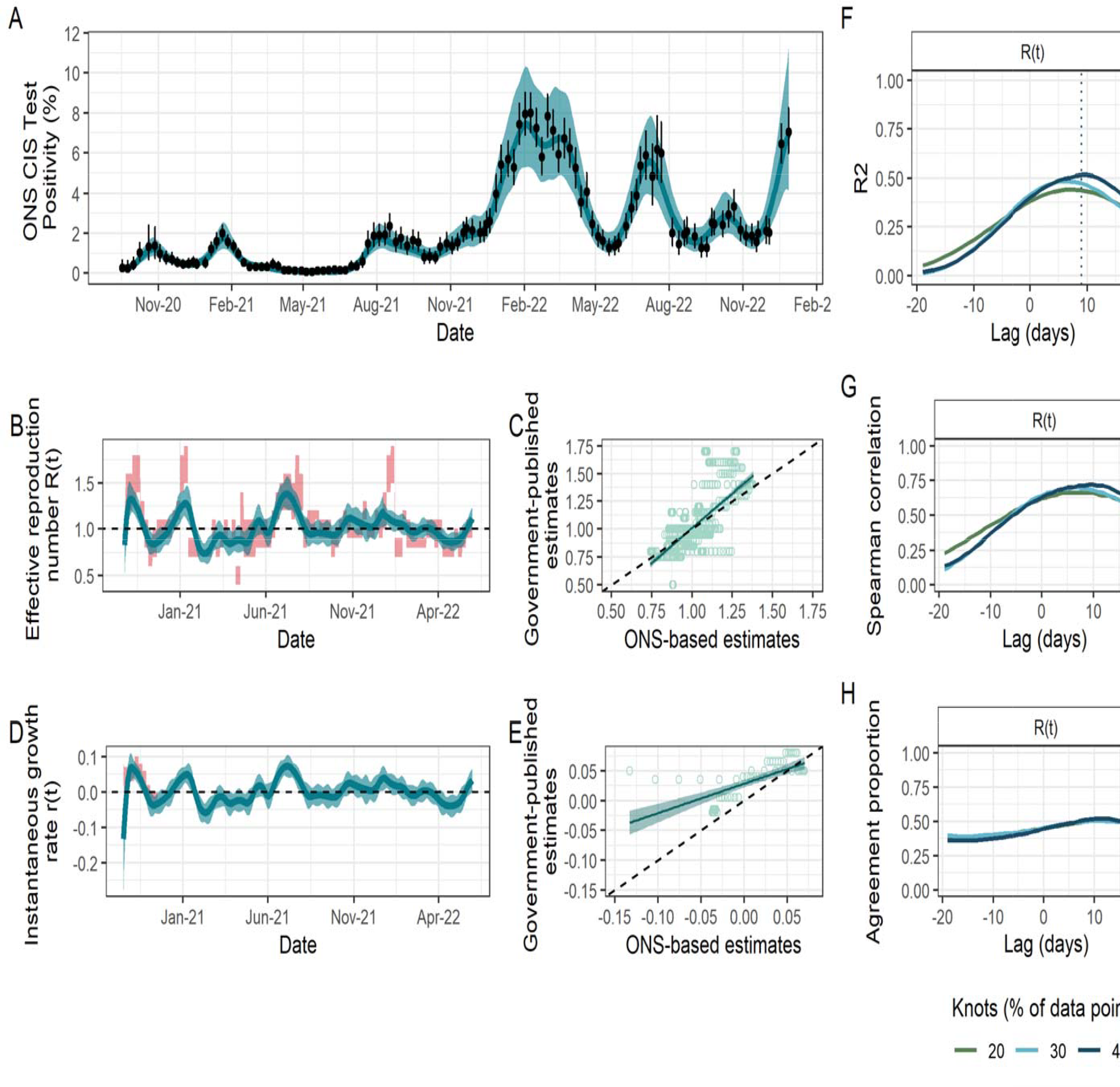
Fit to ONS CIS data, resulting estimates of R(t) and r(t), and comparison metrics for Northern Ireland. (A) Spline model fit 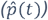 (blue; median line with 95% credible intervals) to data (black points; ONS point estimate with 95% credible intervals) with the number of knots totalling 40% of the total data points. (B) The ONS-based estimated effective reproduction number 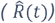 (blue; median line with 95% credible intervals) alongside the government-published estimates 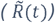 (red; 90% confidence intervals), lagged by 9 days. The black dashed line highlights the epidemic growth threshold of 1. (C) Scatterplot (light green points) showing the relationship between the ONS-based and government-reported estimates of R(t), with the latter lagged by 9 days. The dark green line shows the trend line from the linear model (with dark green shading corresponding to 95% confidence intervals) regressing the lagged government-published estimates on the ONS-based estimates. Rows of points are due to the limited precision available (due to rounding before publication) for government-published estimates. (D) The ONS-based estimated instantaneous growth rate 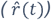 (blue; median line with 95% credible intervals) alongside the government-published estimates 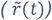 (red; 90% confidence intervals), without any lag. The dashed line highlights the epidemic growth threshold of 0. (E) Scatterplot (light green points) showing the relationship between the ONS-based and government-reported estimates of r(t), without any lag. The dark green line shows the trend line from the linear model (with dark green shading corresponding to 95% confidence intervals) regressing the lagged government-published estimates on the ONS-based estimates. Rows of points are due to the limited precision available (due to rounding before publication) for government-published estimates. (F) – (H) Results of the three metrics used to assess the agreement between ONS-based and government-published estimates of R(t) under different lags and for models fitted with a different number of knots, taken as a percentage of the total data points. (F) R^2^. Dotted lines indicate the lag for which this metric is maximised for R(t) and is thus what is applied to the government-based estimates in panels (B) – (C). (G) Spearman rank correlation. (H) Agreement proportion.

We found that the model in England produced similar values of the three comparison metrics for all numbers of knots considered (Figure 1F-G). However, this was not the case in Scotland and Wales, where the number of knots played a more important role (Figure 2F-G; Figure 3F-G): for example, the maximum observed *R*^2^ for *R*(*t*) in Scotland rose by more than 40% from 0.53 to 0.74 when doubling the number of knots from 20% to 40% of total data points. In addition to requiring a greater number of knots, the spline fits in Scotland and Wales, and additionally in Northern Ireland, all have substantially greater uncertainty which is propagated through to 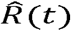 and 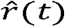. This is likely driven by the increased uncertainty arising from smaller sample sizes in the ONS CIS, which are proportional to the smaller population sizes in these nations compared to England.

Our sensitivity analysis has highlighted a time delay between the ONS-based estimates compared to the government-based estimates. In England, Scotland and Wales, the ONS-based estimates are correlated with later government-published estimates, suggesting that the ONS-based estimates can capture epidemic trends more quickly. (This effect can be seen clearly in Supplementary Feigures 7 – 9). The models in England and Scotland indicated a similar time delay of around 8 days for *R*(*t*) and 10 days for *r*(*t*). In Scotland, *R*^2^ rose by almost 50% from 0.49 with no time delay to its maximum value (0.74) under a delay of 8 days for the *R*(*t*) under the model with 40% knots, emphasising the importance of considering such delays. The time delay which maximised the metrics of interest was much greater in Wales, sitting at around 2 weeks for both parameters.

For *R*(*t*) and *r*(*t*) in England and Scotland, and *R*(*t*) in Wales, the agreement proportion sits close to 0.80 depending on the time delay and number of knots used (range 0.79 – 0.82). The majority of instances in which there is not agreement on whether the epidemic is growing or shrinking occurs for values close to the threshold (e.g. one estimate being slightly over the threshold while the other is slightly under and vice versa), rather than there being large differences between the two estimates. The is what drives the low agreement proportion of *r*(*t*) in Wales, despite the corresponding *R*^2^ and Spearman correlation values being relatively high.

Our ONS-based estimates for Northern Ireland have the weakest relationship with the government-published estimates. 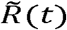 fluctuates substantially more so than 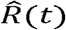 throughout the period, in particular from November 2021 (around the time of the emergence of the Omicron variant). This relationship is reflected in all three comparison metrics, with a maximum of only 50% of the variance in the government-published estimates being explained by the ONS-based estimates. Furthermore, the ONS-based estimates are (weakly) correlated with an earlier government-based, in contrast to England, Scotland and Wales, meaning that in Northern Ireland the ONS-based estimates are slower to track the epidemic trends in this setting.

### Estimates for 2023

Figure 5 presents the spline fits and estimates of 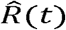 and 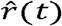 from January – March 2023 in each nation, under varying numbers of knots in the spline model.

**Figure 5:**
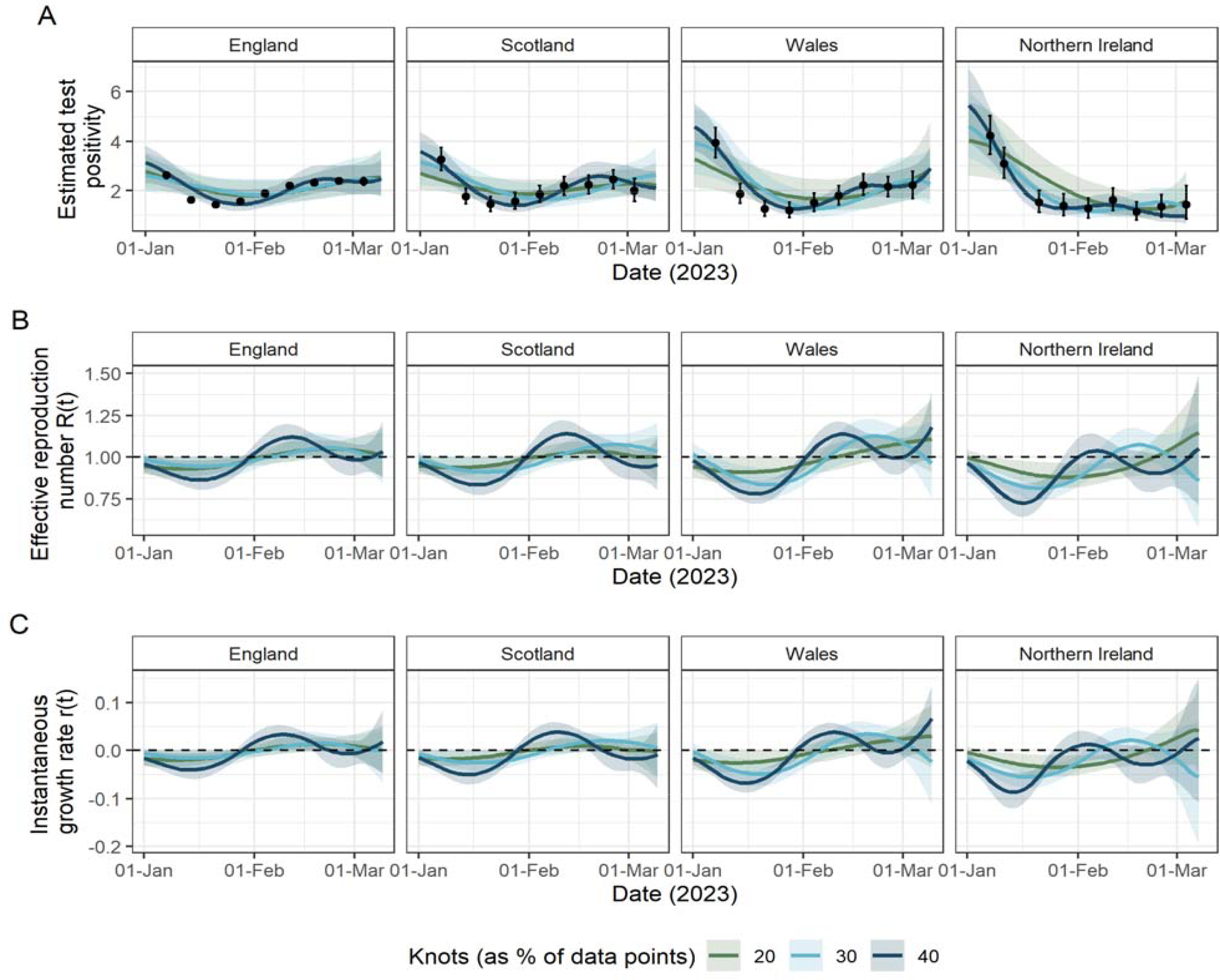
Fit to ONS CIS data and resulting estimates of and for England, Scotland, Wales and Northern Ireland from January 2023 - March 2023, at which point the ONS CIS was initially “paused”. Throughout, lines represent median values and shaded areas are the corresponding 95% credible intervals (A) Spline model fits () to data (black points; ONS point estimate with 95% credible intervals) with the number of knots totalling 20% (green), 30% (light blue) and 40% (purple) of the total data points. (B) The estimated effective reproduction number () resulting from the spline fits in (A). The dashed line highlights the epidemic growth threshold of 1. (C) The estimated instantaneous growth rate () resulting from the spline fits in (A). The dashed line highlights the epidemic growth threshold of 0.

In each nation, ONS test positivity decreases to mid-January, which results in the corresponding estimates falling below the epidemic growth thresholds: 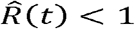 and 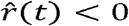. However, the degree to which the estimates indicate a shrinking epidemic is dependent on the number of knots used, with a lower number of knots resulting in the estimates being closer to the threshold and vice versa.

From mid-January, test positivity increases in all nations, peaking in mid-February in England, Scotland and Wales, and in early February in Northern Ireland. This results in epidemic growth, 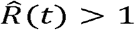 and 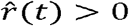, for most of the month of February. In England and Scotland, by the end of the period, both 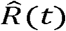 and 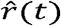 approach the epidemic threshold showing a stable epidemic, but in Wales and Northern Ireland the estimates imply that the epidemic was continuing to grow.

## Discussion

We have demonstrated and validated a method to estimate the *R*(*t*) and *r*(*t*) of SARS-CoV-2 using the publicly available ONS CIS data, which became a primary source characterising the ongoing epidemics in the four nations of the UK after the scaling down of community testing in Spring 2022, until the survey’s initial “pause” in March 2023. We have shown strong agreement between our ONS-based and government-published estimates across mid-2020 until the end of 2022, showing the suitability of this model applied to these data to track the trends in these key epidemic parameters. Specifically, we demonstrated that up to 77% of the variation in the government-published estimates could be explained by our ONS-based estimates, depending on the nation and parameter under consideration, which was complemented by high values of the Spearman rank correlation and agreement proportion. We have also found that most estimates under this model, except for Northern Ireland, led government-reported estimates by up to 2 weeks, a potentially advantageous gap in terms of producing timely real-time modelling estimates. These results are important for the WCIS announced in October 2023,^24^ as the methodology deployed here could potentially be used on the data generated by this study to track community transmission in the UK over winter 2023/24.

Our work is not intended as an alternative to ensemble modelling, which is advantageous in its ability to synthesise information from multiple sources.^43^ Although deployed in multiple different fields,^44–46^ this is particularly important in the context of modelling the SARS-CoV-2 epidemic in the UK, due to the diversity of surveillance data streams available. As discussed by Park et al.^5^, the ensemble modelling approach has many strengths including increased prediction ability and greater robustness. We see our work completing this approach, by presenting a potential solution to the challenge faced by public health officials in the UK in early 2022 given the large scaling-down of the surveillance systems at the time. With less data available, some ensemble models could become less reliable, while the approach presented here would not have been affected given the continuation of the ONS CIS beyond this period.

One of the strengths of this study is its application to all nations of the UK separately, which is uncommon in the literature^40,47–50^. This work underlines the importance of setting-specific analysis of an epidemic and the differences that can arise even when these settings are geographically close and socioeconomically similar. While we were able to produce estimates which matched government-reported estimates closely, we found that both the smoothness of the spline and the time delay between the ONS-based and government-published estimates were dependent on the nation of the UK, with smaller ONS CIS sample sizes (proportional to population size) resulting in less smooth fits. As previously mentioned, this is likely attributable to the increased variability observed in the ONS CIS data among nations with smaller sample sizes. Moreover, each of the four nations have devolved governments which implemented slightly different public health and social measures at varying time points. This nuance would be difficult to capture accurately in a UK-level model.

The importance of setting-specific modelling was underlined in Northern Ireland. We found that, here, 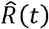 has the weakest relationship with 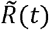 of all of the nations, and this relationship could not even be evaluated for *r*(*t*) due to the lack of government-published estimates in this nation. In addition to the weaker relationship, the temporal relationship between the two sets of estimates were the opposite of that observed in the other three nations, with 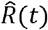 trailing 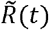. Northern Ireland has the smallest population size of the four nations of the UK (approximately 1.9 million^51^), which inevitably increases the volatility of all surveillance data observed in this setting, and indeed this was demonstrated through the volatility of 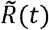. In particular, this volatility may have contributed to the lack of published estimates 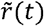, occurring when a consensus estimate from the ensemble models could not be generated due to the small number of reliable model outputs available.

Our model builds upon the methodology set out in ^25^ by adapting the model to fit to ONS CIS data. Of course, other methods also exist to estimate *r*(*t*) and *R*(*t*) using the ONS CIS data. While theoretically it would be possible to use ONS-estimated incidence of infection within previously published frameworks such as ^6,7,52^, these figures were published up to 3 weeks later than the corresponding test positivity estimates and ceased to be published in June 2022, thus making these data unsuitable for ongoing real-time modelling of the epidemic. As an alternative, Abbott and Funk^38^ deconvolve ONS test positivity data into incidence, which they then model using a Gaussian process. However, in addition to the assumption of the generation time distribution, which is a necessity of the renewal model, the probability density function of the time from infection until PCR positivity is also required. Similarly, the ONS CIS is among the data streams that Birrell et al.^39,40^ fit to as part of their age- and NHS-England-region-stratified Susceptible-Exposed-Infected-Recovered (SEIR) model. However, this model is specific to England, and requires additional parameters to calibrate the complex mechanistic model to the observed data, as is the case with mathematical models of this structure. By contrast, our method only has two key parameters which require tuning to the setting of interest. Firstly, the generation time distribution, which is a common assumption in models with an element of mechanistic transmission.^6,7,52^ Secondly, the smoothness of the spline, controlled by the number of knots. For the latter, we conducted a sensitivity analysis to assess the impact of the number of knots and show that there are often multiple viable options, although we found that data sets with smaller sample sizes were better fitted by models with more knots. Other strengths of our model include the ease with which the generation time distribution can be updated over time, as was required due to the emergence of new variants,^33^ and that we only fit to publicly available data, meaning that anyone may reproduce our results and/or adapt the model to similar data in other contexts, for example in different countries or to different pathogens, or even to other data streams if appropriate.

It is important to be aware of the limitations of this work. First, our estimates suffer from boundary effects attributable to the ONS CIS survey starting mid-way through a wave of infection. This is particularly noticeable in Wales and Northern Ireland (also with smaller sample sizes) and may contribute to the lower levels of agreement observed between the ONS-based and government-published estimates for each comparison metric. Second, as discussed, there is not a standard way to select the optimal level of smoothing and thus was determined in this instance via sensitivity analysis. Eales et al.^25^ have taken a similar approach in their selection the number of knots. Third, as *R*(*t*) and *r*(*t*) are not directly observable quantities, there are no “true” values by which to validate our model. Instead, we have focussed on comparison with the government-published estimates and considered the amount of information that the ONS-based estimates alone could yield. This implicitly treats the government-published estimates as reliable and accurate estimates of the true values of *R*(*t*) and *r*(*t*). Fourth, although our results suggest that our ONS-based estimates provide more timely insights into epidemic trends than the corresponding government-based estimates, this does not consider the time delays between the collection and publication of the ONS CIS data. Although this delay would not change the date on which trends occur, it would change the date when the trends would be able to be estimated and so real-time modelling gains may not be fully realised. This could be overcome by more timely access to the ONS CIS data, but this may compromise the data being publicly available and could also make estimation more logistically challenging. Although not a limitation of our work per se, it is also important to highlight that the ONS CIS was expensive to run (approximately £945m to the end of December 2022^53^), which undoubtedly raises questions regarding the long-term implementation of similar schemes in the future.

In this study, we have demonstrated the utility and validity of this model for estimating () and r () using the ONS CIS test positivity data, which was a primary source measuring the ongoing epidemics in the four nations of the UK. Our model provides a reliable means by which to track the ongoing epidemics in each of the four nations of the UK after the scaling down of SARS-CoV-2 surveillance, which was largely reduced due to the transition from “emergency” to “endemic” state in Spring 2022. Although the ONS CIS was “paused” in March 2023, the WCIS (announced in October 2023) should provide a new surveillance data stream, similar to the ONS CIS data, for the winter period 2023/24. Consequently, this model is uniquely placed to provide a reliable method for tracking the spread of the UK epidemic throughout this potentially challenging winter period.

## Data Availability

All data and code used are available from: https://github.com/ruthmccabe/ons-test-positivity-model.

## Acknowledgements

This work was supported by the NIHR HPRU in Emerging and Zoonotic Infections, a partnership between PHE, University of Oxford, University of Liverpool and Liverpool School of Tropical Medicine [grant number NIHR200907 supporting RM and CAD]; and the MRC Centre for Global Infectious Disease Analysis [grant number MR/R015600/1], which is jointly funded by the UK Medical Research Council (MRC) and the UK Foreign, Commonwealth & Development Office (FCDO), under the MRC/FCDO Concordat agreement and is also part of the EDCTP2 programme supported by the European Union (EU). RM was also supported by the Isaac Newton Institute (INI) Knowledge Transfer Network (KTN) in funding a 3-month placement at the UK Health Security Agency.

## Disclaimer

“The views expressed are those of the authors and not necessarily those of the United Kingdom (UK) Department of Health and Social Care, EU, FCDO, MRC, National Health Service, NIHR, PHE or UKHSA. The funding bodies had no role in the design of the study, analysis and interpretation of data and in writing the manuscript.”

## Author contributions

RM conceived the idea for the study following discussions with GD, JP, JPG and CAD. RM conducted the analysis and drafted the manuscript, with all authors contributing to redrafting.

## Data availability

All data and code used are available from: https://github.com/ruthmccabe/ons-test-positivity-model.

## Notes

### Competing Interest Statement

The authors have declared no competing interest.

## References

1. Slides to accompany coronavirus press conference: 11 May 2020. gov.uk https://assets.publishing.service.gov.uk/government/uploads/system/uploads/attachment_data/file/884352/slides_-_11_05_2020.pdf (2020).

2. Slides and datasets to accompany coronavirus press conference: 22 October 2020. gov.uk https://assets.publishing.service.gov.uk/government/uploads/system/uploads/attachment_data/file/928812/Press_conference_slides_-_Thursday_22_October_.pdf (2020).

3. Government publishes latest R number. gov.uk https://www.gov.uk/government/news/government-publishes-latest-r-number (2020).

4. Anderson, R. et al. Reproduction number (R) and growth rate (r) of the COVID-19 epidemic in the UK: methods of estimation, data sources, causes of heterogeneity, and use as a guide in policy formulation. The Royal Society (2020).

5. Park, J. et al. Combining models to generate a consensus effective reproduction number R for the COVID-19 epidemic status in England. medRxiv (2023) doi:10.1101/2023.02.27.23286501.

6. Cori, A., Ferguson, N. M., Fraser, C. & Cauchemez, S. A New Framework and Software to Estimate Time-Varying Reproduction Numbers During Epidemics. Am. J. Epidemiol. 178, 1505–1512 (2013).

7. Parag, K. V. Improved estimation of time-varying reproduction numbers at low case incidence and between epidemic waves. PLoS Comput. Biol. 17, e1009347 (2021).

8. Moore, R. E., Rosato, C. & Maskell, S. Refining epidemiological forecasts with simple scoring rules. Philos. Trans. R. Soc. A Math. Phys. Eng. Sci. 380, 20210305 (2022).

9. Vöhringer, H. S. et al. Genomic reconstruction of the SARS-CoV-2 epidemic in England. Nature 600, 506–511 (2021).

10. Dean, N. E. Tracking COVID-19 infections: time for change. Nature 602, (2022).

11. Clarke, J., Beaney, T. & Majeed, A. UK scales back routine covid-19 surveillance. BMJ 376, o562 (2022).

12. Tapper, J. Dismay as funding for UK’s ‘world-beating’ Covid trackers is axed. The Observer (2022).

13. Coronavirus (COVID-19) in the UK. gov.uk https://coronavirus.data.gov.uk/ (2023).

14. Coronavirus (COVID-19) Infection Survey, UK. Office for National Statistics https://www.ons.gov.uk/surveys/informationforhouseholdsandindividuals/householdan dindividualsurveys/covid19infectionsurvey (2022).

15. Real-time Assessment of Community Transmission (REACT) Study. Imperial College London https://www.imperial.ac.uk/medicine/research-and-impact/groups/reactstudy/ (2022).

16. The COVID-19 Genomics UK (COG-UK) consortium. An integrated national scale SARS-CoV-2 genomic surveillance network. The Lancet Microbe 1, e99–e100 (2020).

17. COVID–19 Genomic Surveillance. Wellcome Sanger Institute https://covid19.sanger.ac.uk/lineages/raw (2023).

18. Morvan, M. et al. An analysis of 45 large-scale wastewater sites in England to estimate SARS-CoV-2 community prevalence. Nat. Commun. 13, 4313 (2022).

19. COVID-19 Response: Living with COVID-19. Cabinet Office https://www.gov.uk/government/publications/covid-19-response-living-with-covid-19/covid-19-response-living-with-covid-19 (2022).

20. Coronavirus (COVID-19): Test and Protect - transition plan. Scottish Government https://www.gov.scot/publications/test-protect-transition-plan/ (2022).

21. Together for a safer future: Wales’ long-term Covid-19 transition from pandemic to endemic. Welsh Government https://gov.wales/sites/default/files/publications/2022-05/wales-long-term-covid-19-transition-from-pandemic-to-endemic.pdf (2022).

22. COVID-19 Test, Trace and Protect Transition Plan. Department of Health, Northern Ireland https://www.health-ni.gov.uk/sites/default/files/publications/health/Test-Trace-Transition-Plan.pdf (2022).

23. COVID-19 Infection Survey participants thanked for ‘huge contribution’ to pandemic response. UK Health Security Agency https://www.gov.uk/government/news/covid-19-infection-survey-participants-thanked-for-huge-contribution-to-pandemic-response (2023).

24. UKHSA and ONS launch new Winter COVID-19 Infection Study. UK Health Security Agency https://ukhsa-newsroom.prgloo.com/news/ukhsa-and-ons-launch-new-winter-covid-19-infection-study (2023).

25. Eales, O. et al. Appropriately smoothing prevalence data to inform estimates of growth rate and reproduction number. Epidemics 40, (2022).

26. Coronavirus (COVID-19) Infection Survey: methods and further information. Office for National Statistics https://www.ons.gov.uk/peoplepopulationandcommunity/healthandsocialcare/conditionsanddiseases/methodologies/covid19infectionsurveypilotmethodsandfurtherinformati on (2023).

27. Coronavirus (COVID-19) Infection Survey, UK: 15 July 2022. Office for National Statistics https://www.ons.gov.uk/peoplepopulationandcommunity/healthandsocialcare/conditionsanddiseases/bulletins/coronaviruscovid19infectionsurveypilot/15july2022 (2022).

28. Coronavirus (COVID-19) Infection Survey, UK: 24 March 2023. Office for National Statistics https://www.ons.gov.uk/peoplepopulationandcommunity/healthandsocialcare/conditionsanddiseases/bulletins/coronaviruscovid19infectionsurveypilot/24march2023 (2023).

29. Ward, H. et al. SARS-CoV-2 antibody prevalence in England following the first peak of the pandemic. Nat. Commun. 12, 905 (2021).

30. Wallinga, J. & Lipsitch, M. How generation intervals shape the relationship between growth rates and reproductive numbers. Proceedings. Biol. Sci. 274, 599–604 (2007).

31. Homan, M. D. & Gelman, A. The No-U-Turn Sampler: Adaptively Setting Path Lengths in Hamiltonian Monte Carlo. J. Mach. Learn. Res. 15, 1593–1623 (2014).

32. Wallinga, J. & Teunis, P. Different Epidemic Curves for Severe Acute Respiratory Syndrome Reveal Similar Impacts of Control Measures. Am. J. Epidemiol. 160, 509–516 (2004).

33. Hart, W. S. et al. Generation time of the alpha and delta SARS-CoV-2 variants: an epidemiological analysis. Lancet Infect. Dis. 22, 603–610 (2022).

34. SARS-CoV-2 variants in analyzed sequences, United Kingdom. Our World in Data https://ourworldindata.org/grapher/covid-variants-area?country=~GBR (2023).

35. Hart, W. S. et al. Inference of the SARS-CoV-2 generation time using UK household data. Elife 11, e70767 (2022).

36. Abbott, S., Sherratt, K., Gerstung, M. & Funk, S. Estimation of the test to test distribution as a proxy for generation interval distribution for the Omicron variant in England. medRxiv 2022.01.08.22268920 (2022) doi:10.1101/2022.01.08.22268920.

37. Reproduction number (R) and growth rate: methodology. gov.uk https://www.gov.uk/government/publications/reproduction-number-r-and-growth-rate-methodology/reproduction-number-r-and-growth-rate-methodology (2021).

38. Abbott, S. & Funk, S. Estimating epidemiological quantities from repeated cross-sectional prevalence measurements. medRxiv (2022) doi:10.1101/2022.03.29.22273101.

39. Birrell, P., Blake, J., Leeuwen, E. van Angelis., D. De & MRC Biostatistics Unit COVID-19 Working Group. COVID-19: nowcast and forecast. MRC Biostatistics Unit https://joshuablake.co.uk/public-RTM-reports/iframe.html (2022).

40. Birrell, P., Blake, J., van Leeuwen, E., Gent, N. & De Angelis, D. Real-time nowcasting and forecasting of COVID-19 dynamics in England: the first wave. Philos. Trans. R. Soc. B Biol. Sci. 376, 20200279 (2021).

41. Maishman, T. et al. Statistical methods used to combine the effective reproduction number, R(t), and other related measures of COVID-19 in the UK. Stat. Methods Med. Res. 31, 1757–1777 (2022).

42. The R value and growth rate. gov.uk https://www.gov.uk/guidance/the-r-value-and-growth-rate (2022).

43. Barai, S. V & Reich, Y. Ensemble modelling or selecting the best model: Many could be better than one. AI EDAM 13, 377–386 (1999).

44. Di Napoli, M. et al. Machine learning ensemble modelling as a tool to improve landslide susceptibility mapping reliability. Landslides 17, 1897–1914 (2020).

45. Grenouillet, G., Buisson, L., Casajus, N. & Lek, S. Ensemble modelling of species distribution: the effects of geographical and environmental ranges. Ecography (Cop.). 34, 9–17 (2011).

46. Zhou, L., Lai, K. K. & Yu, L. Least squares support vector machines ensemble models for credit scoring. Expert Syst. Appl. 37, 127–133 (2010).

47. Danon, L., Brooks-Pollock, E., Bailey, M. & Keeling, M. A spatial model of COVID-19 transmission in England and Wales: early spread, peak timing and the impact of seasonality. Philos. Trans. R. Soc. B Biol. Sci. 376, 20200272 (2021).

48. Simpson, C. R. et al. Temporal trends and forecasting of COVID-19 hospitalisations and deaths in Scotland using a national real-time patient-level data platform: a statistical modelling study. Lancet Digit. Heal. 3, e517–e525 (2021).

49. Keeling, M. J. et al. Fitting to the UK COVID-19 outbreak, short-term forecasts and estimating the reproductive number. Stat. Methods Med. Res. 31, 1716–1737 (2022).

50. Davies, N. G., Kucharski, A. J., Eggo, R. M., Gimma, A. & Edmunds, W. J. Effects of non-pharmaceutical interventions on COVID-19 cases, deaths, and demand for hospital services in the UK: a modelling study. Lancet. Public Heal. 5, e375–e385 (2020).

51. Northern Ireland population mid-year estimate. Office for National Statistics https://www.ons.gov.uk/peoplepopulationandcommunity/populationandmigration/populationestimates/timeseries/nipop/pop (2022).

52. Scott, J. A. et al. Epidemia: An R Package for Semi-Mechanistic Bayesian Modelling of Infectious Diseases using Point Processes. arXiv (2021) 10.48550/arxiv.2110.12461.

53. COVID-19 Infection Survey cost. Office for National Statistics https://www.ons.gov.uk/aboutus/transparencyandgovernance/freedomofinformationfoi/covid19infectionsurveycost (2023).

