## Supplementary Material for "Inferring community transmission of SARS-CoV-2 in the United Kingdom using the ONS COVID-19 Infection Survey"

Ruth McCabe[1,2,3], Gabriel Danelian[3], Jasmina Panovska-Griffiths[3,4,5], Christl A. Donnelly[1,2,5,6]

[1] Department of Statistics, University of Oxford

[2] National Institute for Health and Care Research Health Protection Research Unit in Emerging and Zoonotic Infections

[3] United Kingdom Health Security Agency

[4] The Queen's College, University of Oxford

[5] The Pandemic Sciences Institute, University of Oxford

[6] MRC Centre for Global Infectious Disease Analysis, Imperial College London

### ONS test positivity spline model

#### Definition of B-splines

Given a sequence of non-decreasing knots, $t_1, \dots, t_J$, the $k$th first-order B-spline is defined as:

$$B_{k,1}(t) = \begin{cases} 1 \; if \; t_k < t < t_{k+1} \\ 0 \; otherwise \end{cases}.$$

The second and third order $k$th B-splines are then defined recursively as follows:

$$B_{k,2}(t) = \omega_{k,2} B_{k,1}(t) + \omega_{k,2} B_{k+1,1}(t)$$

$$B_{k,3}(t) = \omega_{k,3} B_{k,2}(t) + \omega_{k,3} B_{k+1,2}(t)$$

where:

$$\omega_{k,2} = \begin{cases} \dfrac{t - t_k}{t_{k+1} - t_k} & if \; t_k \neq t_{k+1} \\ 0 & otherwise \end{cases}$$

$$\omega_{k,3} = \begin{cases} \dfrac{t - t_k}{t_{k+2} - t_k} & if \; t_k \neq t_{k+2} \\ 0 & otherwise \end{cases}.$$

#### Parameterising the Beta distribution

Recall from the main text that:

- $\mu_t$ denotes the central ONS estimate at time $t$;
- $l_t$ and $u_t$ denote the corresponding lower and upper credible intervals, respectively;
- $\sigma_t^2 \approx \left(\frac{u_t - l_t}{3.92}\right)^2$ denotes the variance;
- $\pi_t \sim Beta(\alpha_t, \beta_t)$ is a random variable for the proportion of people testing positive via the ONS survey.

For a random variable distributed according to a Beta distribution, such as $X_t$, it follows that:

$$\mathbb{E}(X_t) = \frac{\alpha_t}{\alpha_t + \beta_t}$$

$$Var(X_t) = \frac{\alpha_t \beta_t}{(\alpha_t + \beta_t)^2 (\alpha_t + \beta_t + 1)}.$$

By setting $\mathbb{E}(X_t) = \mu_t$ and $Var(X_t) = \sigma_t^2$, rearranging provides the following expressions for $\alpha_t$ and $\beta_t$:

$$\alpha_t = \mu_t \left( \frac{\mu_t(1-\mu_t)}{\sigma_t^2} - 1 \right) > 0$$

$$\beta_t = \alpha_t \left( \frac{1-\mu_t}{\mu_t} \right) > 0.$$

### Simulation study to investigate use of Normal likelihood

A simulation study was used in order to examine the appropriateness of using a Normal likelihood distribution in the model. In short, prevalence over time is simulated and by assuming a constant number of tests, the number of positives given the simulated prevalence is determined. The number of positive and total tests can be used to provide a central estimate and 95% confidence interval like the format of the publicly available ONS CIS estimates. We then proceed with the first step of the model as described in the main text, by using the data provided to sample from the resulting Beta distribution (see previous section) and compare the sampled values to a Normal distribution.

Let $\pi_t$ denote the test positivity at time $t$. An iterative random-walk procedure is used to simulate values of $\pi_t$ for $t = 1, \ldots, T$ as follows:

1. Initialise $\pi_1 \sim Beta(2,10)$.
2. To prevent sampling a value of $\pi_t$ which falls out of the interval $[0-1]$, $\pi_1$ is transformed to the logit scale via $logit(\pi_1) = \log\left(\frac{\pi_1}{1-\pi_1}\right)$.
3. For $t = 2, \ldots, T$, $logit(\pi_t) \sim Normal(logit(\pi_{t-1}), \zeta)$, where $\zeta \sim Normal(2,50)$.
4. For all $t$, transform $logit(\pi_t)$ back to the interval $[0-1]$ using $\pi_t = \frac{\exp(logit(\pi_t))}{\exp(logit(\pi_t))+1}$.

Denote by $y_t$ the number of positive tests and by $n_t$ the total number of tests at time $t$. It is assumed that $n_t = n$ is held constant across the period. For each time point, we determine $y_t = \pi_t \times n$.

Exact 95% binomial confidence intervals are obtained using the number of positives and total tests per time point. To match the format of the ONS data, the maximum likelihood estimate (MLE) $\left(\frac{y_t}{n}\right)$ is taken as the central estimate ($\mu_t$) with the 95% confidence intervals as the uncertainty range ($l_t$ and $u_t$).

We assume $n = 80{,}000$ tests are conducted at each time point, in accordance with the number of participants in the ONS CIS in England in December 2022,[1] and we let $T = 150$. This produces data such as that presented in Supplementary Figure 1.

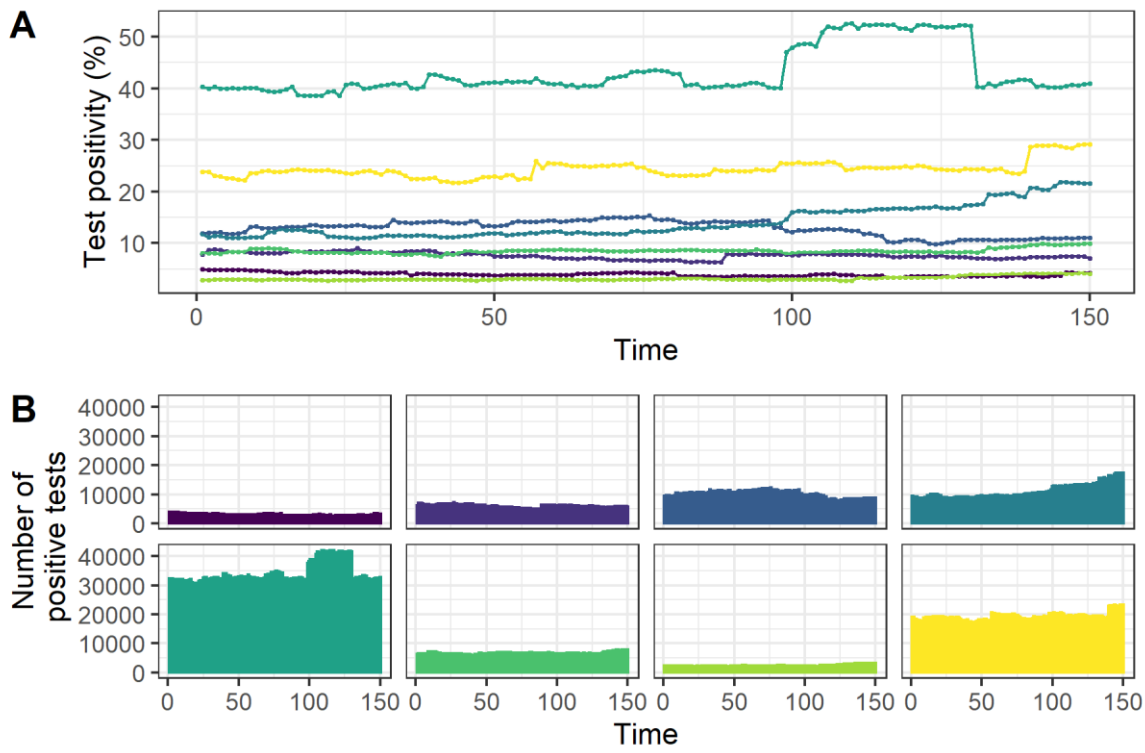

*Supplementary Figure 1: Examples of data simulated to test the assumption of using a Normal likelihood. Each colour represents a distinct simulation and is used to link the outputs in (A) and (B). At each time point of the simulation, it is assumed that $n = 80{,}000$ tests are conducted. (A) Test positivity ($\pi_t$) simulated from the random walk procedure. (B) The number of positive tests determined via $x_t = \pi_t \times n$.*

Having simulated data in the format of the publicly available ONS estimates, we proceed with mimicking the initial sampling phase of the model. Specifically:

$$\pi_t \sim Beta(\alpha_t, \beta_t)$$

where:

$$\alpha_t = \mu_t \left( \frac{\mu_t(1-\mu_t)}{\sigma_t^2} - 1 \right) > 0$$

$$\beta_t = \alpha_t \left( \frac{1-\mu_t}{\mu_t} \right) > 0$$

$$\sigma_t^2 \approx \left( \frac{u_t - l_t}{3.92} \right)^2.$$

For each $t$, we sample from the Beta distribution 10,000 times and take the logit of each. Examples of this for one set of simulated data are shown in Supplementary Figure 2. The resulting (logit) samples are compared to a Normal distribution with mean and standard deviation calculated from the 10,000 samples using a Kolmogorov-Smirnov test for each $t = 1, \dots, 150$.

The entire process of simulating data, sampling 10,000 times from the resulting Beta distribution for each $t = 1, \dots, 150$ and comparing this to Normal distribution using a

Kolmogorov-Smirnov test is repeated 1,000 times to account for the variability arising from the simulation process. Across the 1,000 repetitions, most time points were Normally distributed according to Kolmogorov-Smirnov tests with a critical p-value threshold of $0.05$. 33 of the repetitions (3.3%) had 1 time point out of 150 that was not-normally distributed (0.7%) and 2 repetitions (0.2%) had 2 time points out of 150 that were not normally distributed (1.3%). Consequently, the Normal distribution is deemed appropriate the likelihood in the ONS CIS spline model.

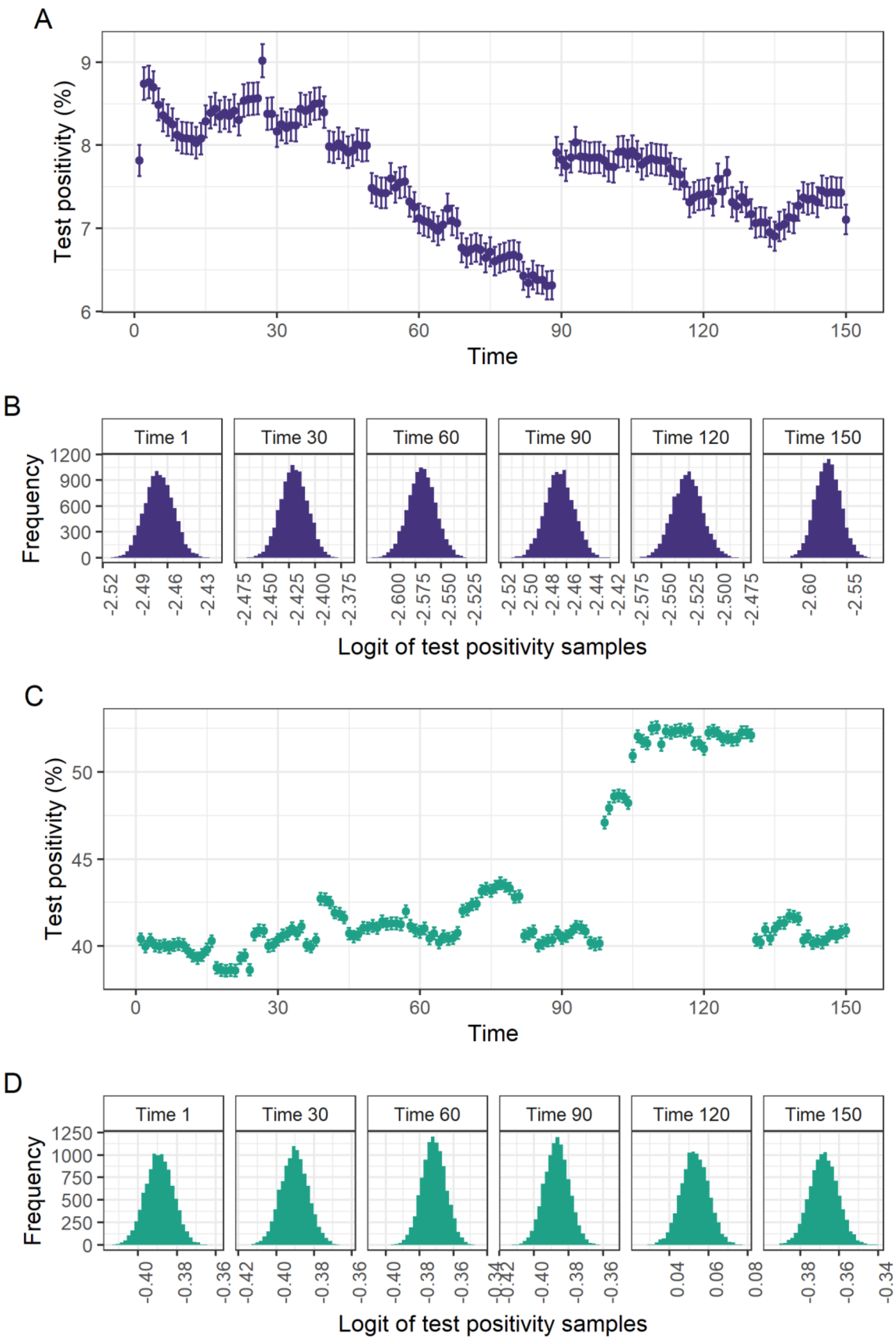

*Supplementary Figure 2: Examples of simulated data and 10,000 samples from resulting Beta distribution at 6 equidistant time periods in the simulation. Data in the two examples here are also shown in Supplementary Figure 1. (A) and (C) show the simulated test positivity time series formatted in the style of the publicly available ONS CIS estimates (central estimate with 95% confidence intervals). (B) and (D) present histograms of the 10,000 samples transformed to the logit scale at 6 time points corresponding to the data shown in (A) and (C), respectively.*

### Diagnostic traceplots

Supplementary Figure 3, Supplementary Figure 4, Supplementary Figure 5 and Supplementary Figure 6 present a selection of traceplots for parameters in the model fit to data from England, Scotland, Wales and Northern Ireland, respectively.

Recall the model specification from the main text:

$$\pi_t \sim Beta(\alpha_t, \beta_t)$$

$$a_1 \sim Normal(0,1)$$

$$a_k \sim Normal(a_{k-1}, \tau^2)$$

$$\tau^2 \sim InverseGamma(0.0001, 0.0001)$$

$$logit(\hat{p}(t)) = \sum_{k=1}^{K} a_k B_{k,3}(t)$$

$$logit(\pi_t) \sim Normal(logit(\hat{p}(t)), \gamma^2)$$

$$\gamma^2 \sim InverseGamma(0.0001, 0.0001)$$

In each plot, $a[k]$ represents $a_k$; $Y\_hat[k]$ represents $\hat{p}(k)$ for discrete time points $k$, which are selected as the first ($k = 1$), the last time, and then equidistant between the first and last time points of the ONS CIS data being fitted to.

$tau$ and $gamma$ represent $\tau$ and $\gamma$ as defined above, respectively.

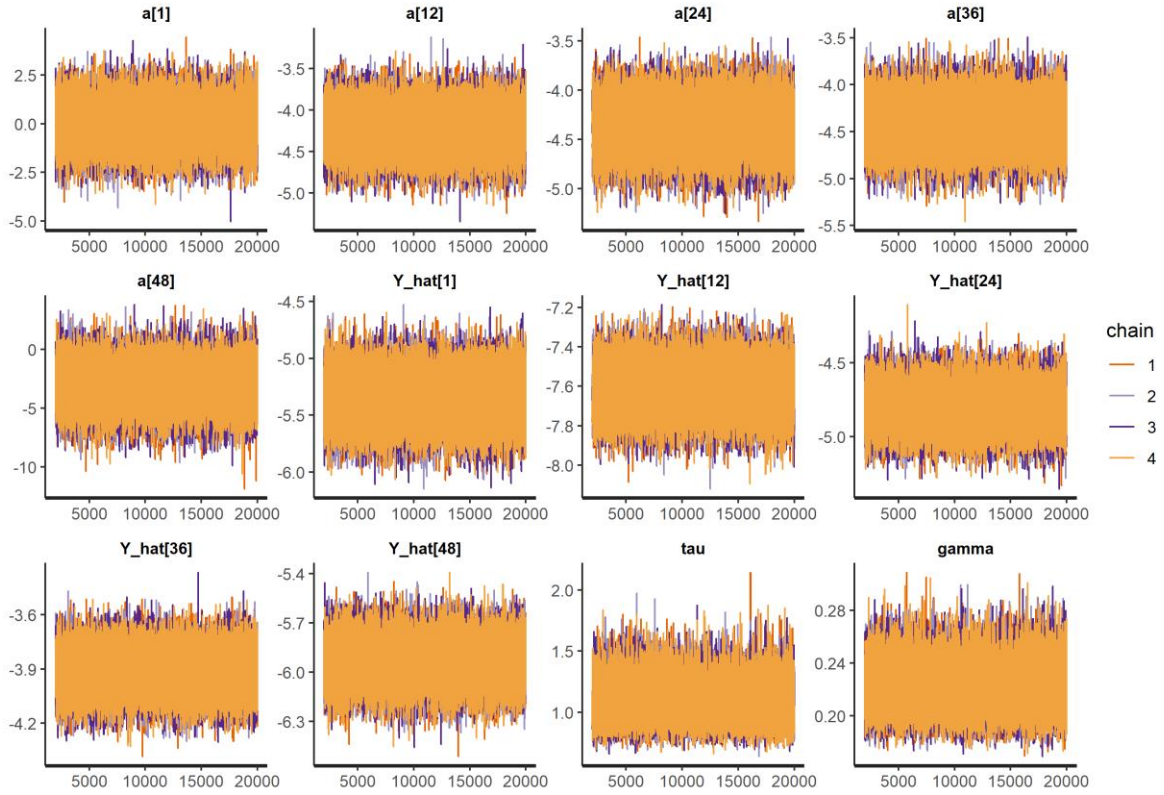

*Supplementary Figure 3: Traceplots for a subset of parameters in the model fit to data from England.*

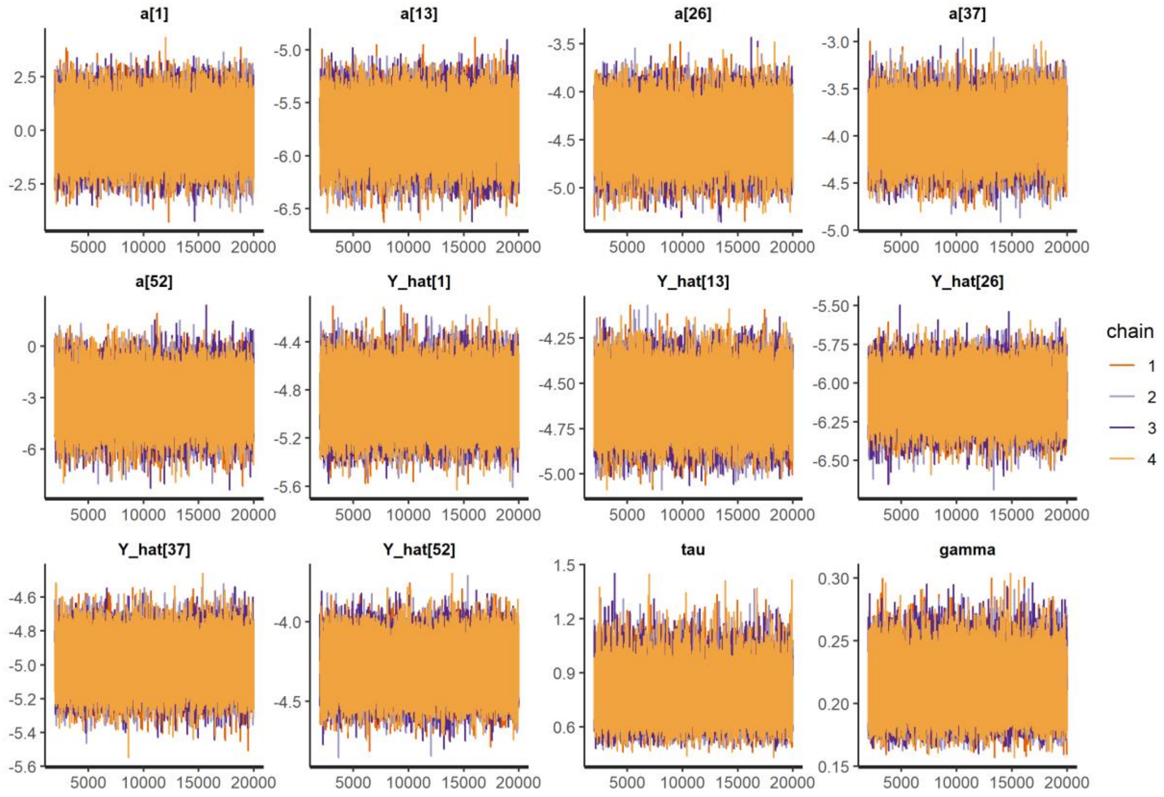

*Supplementary Figure 4: Traceplots for a subset of parameters in the model fit to data from Scotland.*

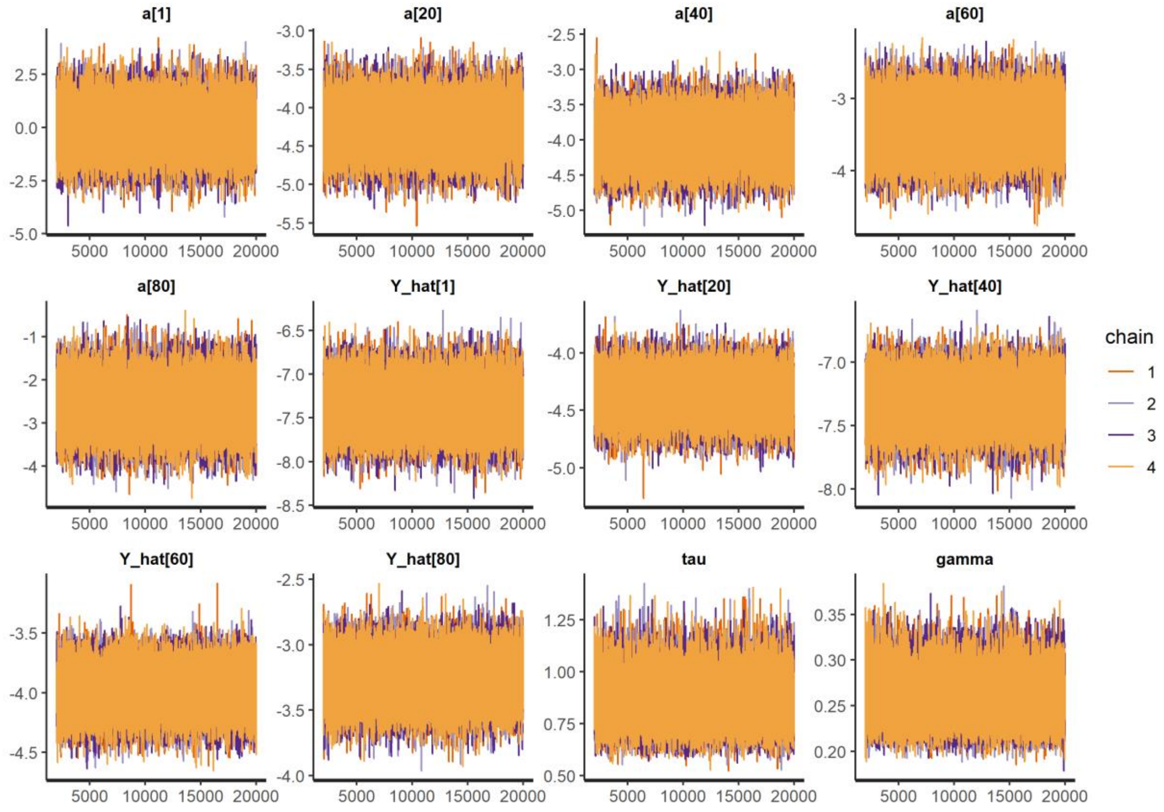

*Supplementary Figure 5: Traceplots for a subset of parameters in the model fit to data from Wales.*

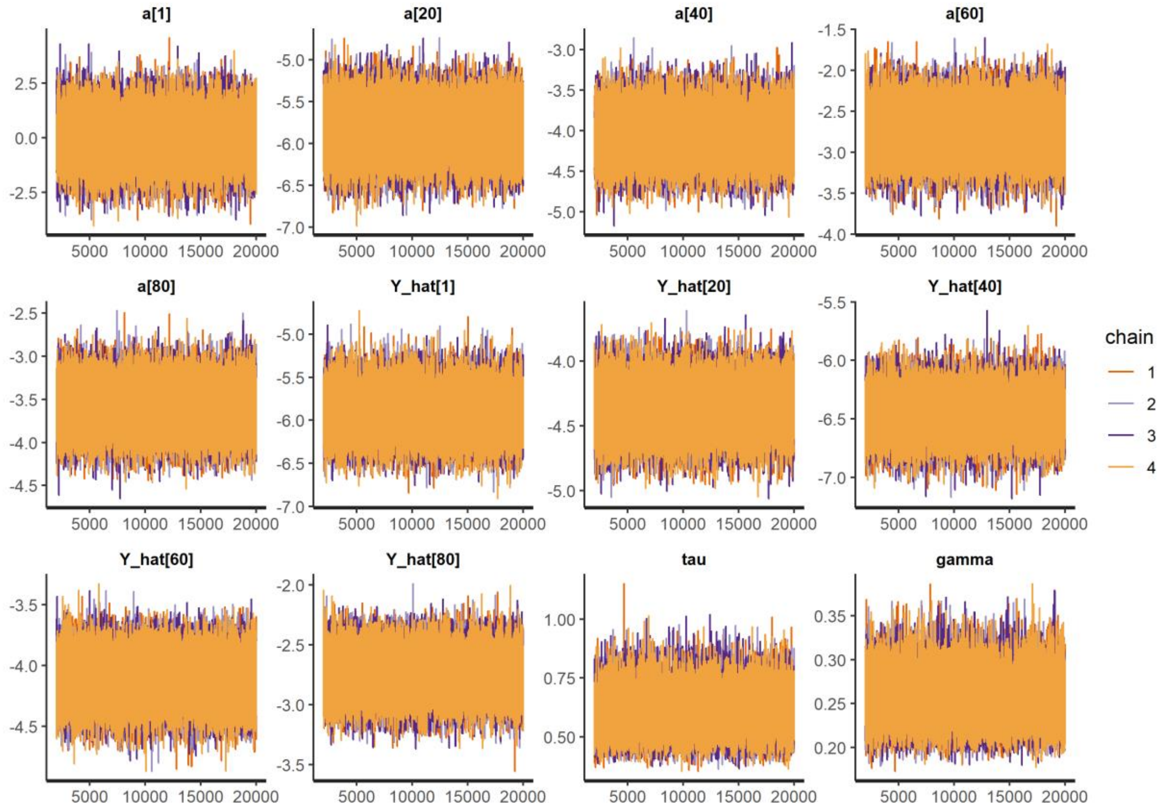

*Supplementary Figure 6: Traceplots for a subset of parameters in the model fit to data from Northern Ireland.*

### Relationship between ONS-based and government-published estimates

#### Comparison of estimates without lags

Supplementary Figure 7, Supplementary Figure 8, Supplementary Figure 9 and Supplementary Figure 10 present the spline fits and resulting estimates $\hat{R}(t)$ and $\hat{r}(t)$ which are compared to $\tilde{R}(t)$ and $\tilde{r}(t)$, respectively, without any time lags.

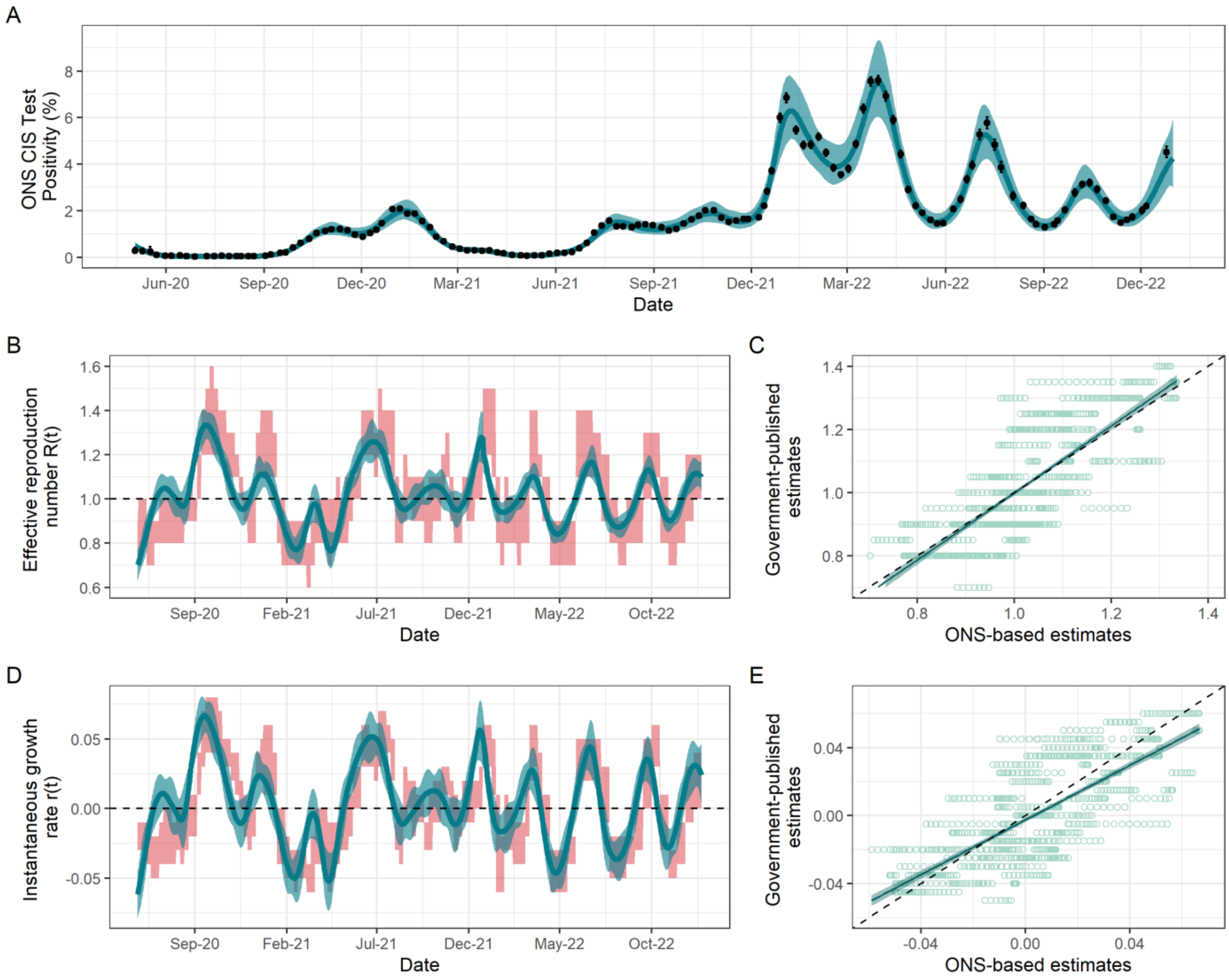

*Supplementary Figure 7: Fit to ONS CIS data and resulting estimates of $R(t)$ and $r(t)$ for England without any lags. (A) Spline model fit ($\hat{p}(t)$) (blue; median line with 95% credible intervals) to data (black points; ONS point estimate with 95% credible intervals) with the number of knots totalling 30% of the total data points. (B) The ONS-based estimated effective reproduction number ( $\hat{R}(t)$) (blue; median line with 95% credible intervals) alongside the government-published estimates ( $\tilde{R}(t)$) (red; 90% confidence intervals). The black dashed line highlights the epidemic growth threshold of 1. (C) Scatterplot (light green points) showing the relationship between the ONS-based and government-reported estimates of $R(t)$. The dark green line shows the trend line from the linear model (with dark green shading corresponding to 95% confidence intervals) regressing the government-published estimates on the ONS-based estimates. Rows of points are due to the limited precision available (due to rounding before publication) for government-published estimates. (D) The ONS-based estimated instantaneous growth rate ( $\hat{r}(t)$) (blue; median line with 95% credible intervals) alongside the government-published estimates ( $\tilde{r}(t)$) (red; 90% confidence intervals). The dashed line highlights the epidemic growth threshold of 0. (E) Scatterplot (light green points) showing the relationship between the ONS-based and government-reported estimates of $r(t)$. The dark green line shows the trend line from the linear model (with dark green shading corresponding to 95% confidence intervals) regressing the lagged government-published estimates on the ONS-based estimates. Rows of points are due to the limited precision available (due to rounding before publication) for government-published estimates.*

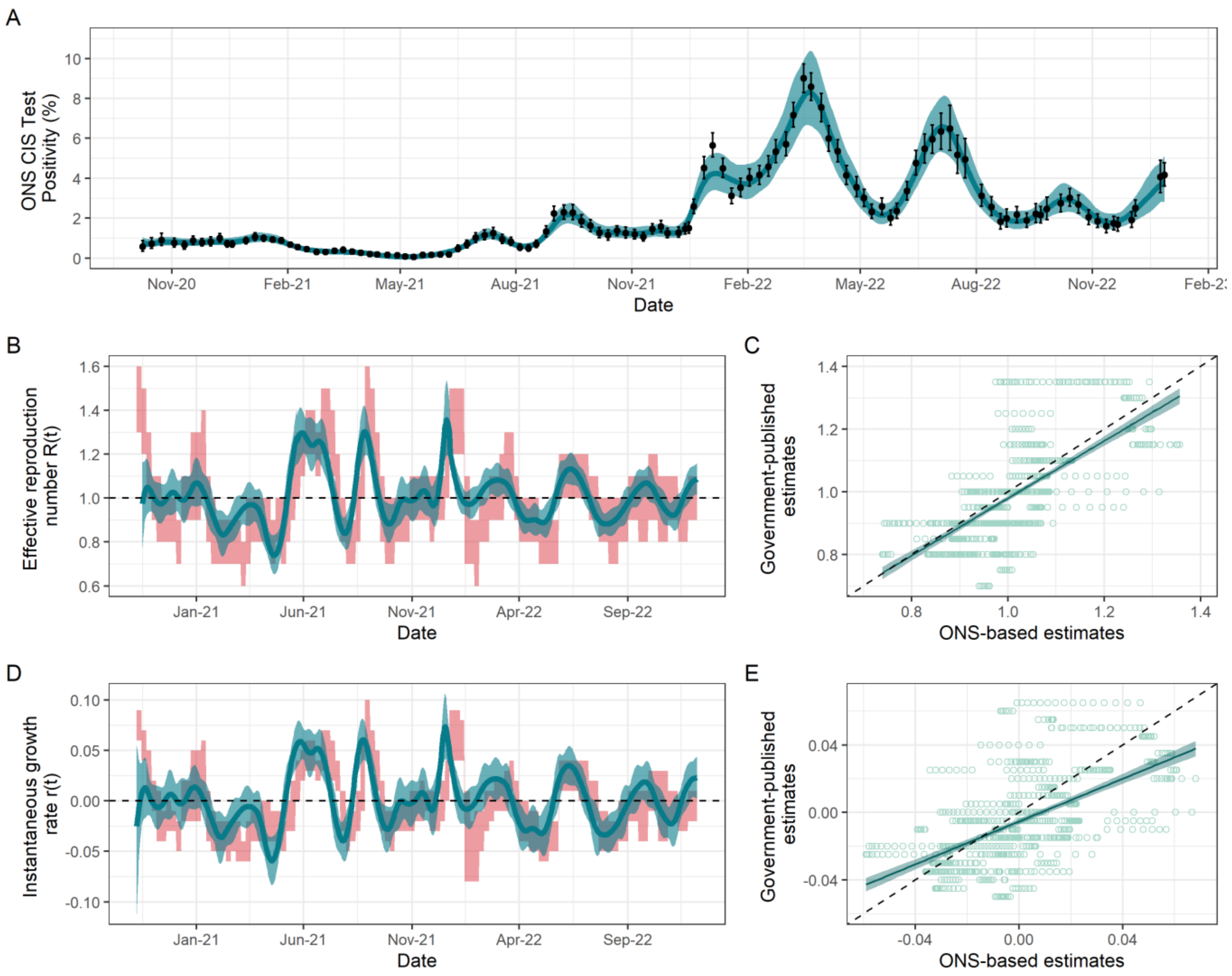

*Supplementary Figure 8: Fit to ONS CIS data and resulting estimates of $R(t)$ and $r(t)$ for Scotland without any lags. (A) Spline model fit ($\hat{p}(t)$) (blue; median line with 95% credible intervals) to data (black points; ONS point estimate with 95% credible intervals) with the number of knots totalling 40% of the total data points. (B) The ONS-based estimated effective reproduction number ( $\hat{R}(t)$) (blue; median line with 95% credible intervals) alongside the government-published estimates ( $\tilde{R}(t)$) (red; 90% confidence intervals). The black dashed line highlights the epidemic growth threshold of 1. (C) Scatterplot (light green points) showing the relationship between the ONS-based and government-reported estimates of $R(t)$. The dark green line shows the trend line from the linear model (with dark green shading corresponding to 95% confidence intervals) regressing the government-published estimates on the ONS-based estimates. Rows of points are due to the limited precision available (due to rounding before publication) for government-published estimates. (D) The ONS-based estimated instantaneous growth rate ( $\hat{r}(t)$) (blue; median line with 95% credible intervals) alongside the government-published estimates ( $\tilde{r}(t)$) (red; 90% confidence intervals). The dashed line highlights the epidemic growth threshold of 0. (E) Scatterplot (light green points) showing the relationship between the ONS-based and government-reported estimates of $r(t)$. The dark green line shows the trend line from the linear model (with dark green shading corresponding to 95% confidence intervals) regressing the lagged government-published estimates on the ONS-based estimates. Rows of points are due to the limited precision available (due to rounding before publication) for government-published estimates.*

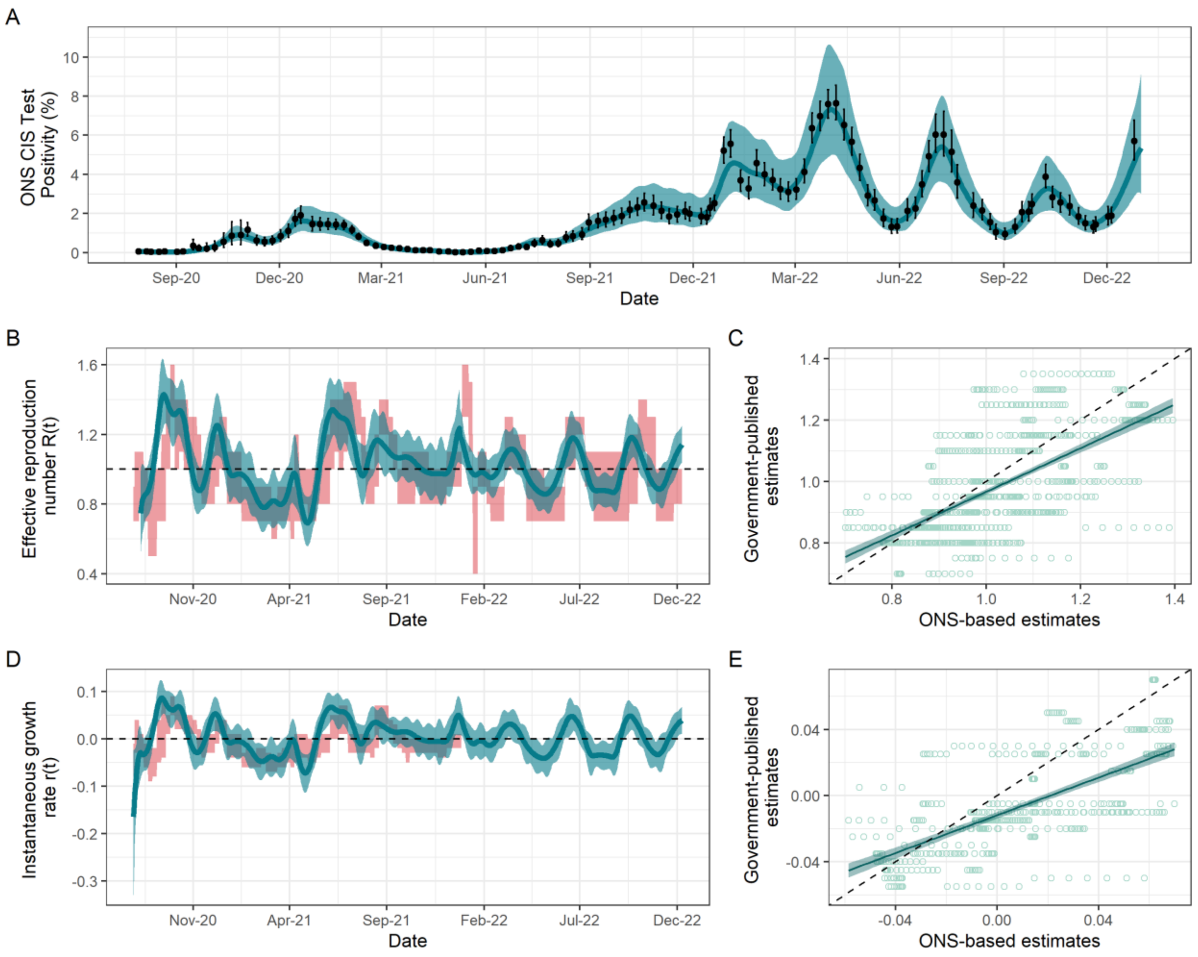

*Supplementary Figure 9: Fit to ONS CIS data and resulting estimates of $R(t)$ and $r(t)$ for Wales without any lags. (A) Spline model fit ($\hat{p}(t)$) (blue; median line with 95% credible intervals) to data (black points; ONS point estimate with 95% credible intervals) with the number of knots totalling 40% of the total data points. (B) The ONS-based estimated effective reproduction number ( $\hat{R}(t)$) (blue; median line with 95% credible intervals) alongside the government-published estimates ( $\tilde{R}(t)$) (red; 90% confidence intervals). The black dashed line highlights the epidemic growth threshold of 1. (C) Scatterplot (light green points) showing the relationship between the ONS-based and government-reported estimates of $R(t)$. The dark green line shows the trend line from the linear model (with dark green shading corresponding to 95% confidence intervals) regressing the government-published estimates on the ONS-based estimates. Rows of points are due to the limited precision available (due to rounding before publication) for government-published estimates. (D) The ONS-based estimated instantaneous growth rate ( $\hat{r}(t)$) (blue; median line with 95% credible intervals) alongside the government-published estimates ( $\tilde{r}(t)$) (red; 90% confidence intervals). The dashed line highlights the epidemic growth threshold of 0. (E) Scatterplot (light green points) showing the relationship between the ONS-based and government-reported estimates of $r(t)$. The dark green line shows the trend line from the linear model (with dark green shading corresponding to 95% confidence intervals) regressing the lagged government-published estimates on the ONS-based estimates. Rows of points are due to the limited precision available (due to rounding before publication) for government-published estimates.*

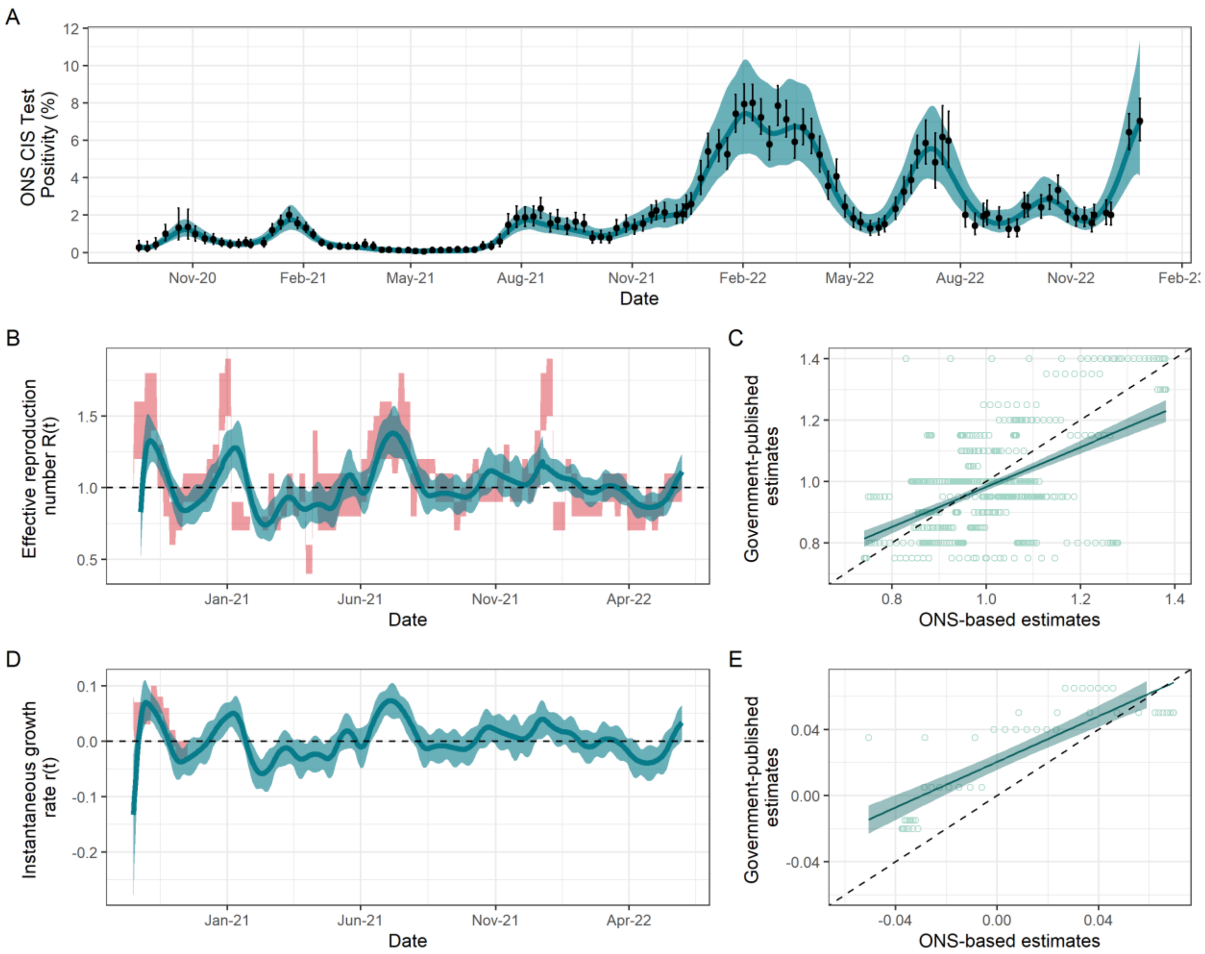

*Supplementary Figure 10: Fit to ONS CIS data and resulting estimates of $R(t)$ and $r(t)$ for Northern Ireland without any lags. (A) Spline model fit ($\hat{p}(t)$) (blue; median line with 95% credible intervals) to data (black points; ONS point estimate with 95% credible intervals) with the number of knots totalling 40% of the total data points. (B) The ONS-based estimated effective reproduction number ( $\hat{R}(t)$) (blue; median line with 95% credible intervals) alongside the government-published estimates ( $\tilde{R}(t)$) (red; 90% confidence intervals). The black dashed line highlights the epidemic growth threshold of 1. (C) Scatterplot (light green points) showing the relationship between the ONS-based and government-reported estimates of $R(t)$. The dark green line shows the trend line from the linear model (with dark green shading corresponding to 95% confidence intervals) regressing the government-published estimates on the ONS-based estimates. Rows of points are due to the limited precision available (due to rounding before publication) for government-published estimates. (D) The ONS-based estimated instantaneous growth rate ( $\hat{r}(t)$) (blue; median line with 95% credible intervals) alongside the government-published estimates ( $\tilde{r}(t)$) (red; 90% confidence intervals). The dashed line highlights the epidemic growth threshold of 0. (E) Scatterplot (light green points) showing the relationship between the ONS-based and government-reported estimates of $r(t)$. The dark green line shows the trend line from the linear model (with dark green shading corresponding to 95% confidence intervals) regressing the lagged government-published estimates on the ONS-based estimates. Rows of points are due to the limited precision available (due to rounding before publication) for government-published estimates.*

### Regression analysis without inclusion of an intercept

In the main text, we set out the following linear regression models to examine the variation of the government-published estimates explained by the ONS-based estimates:

$$\tilde{R}(t) = \beta_0 + \beta_1 \hat{R}(t) + \epsilon$$

and

$$\tilde{r}(t) = \gamma_0 + \gamma_1 \hat{r}(t) + \epsilon.$$

Here, we consider similar models but without the intercept terms $\beta_0$ and $\gamma_0$:

$$\tilde{R}(t) = \beta_1 \hat{R}(t) + \epsilon$$

and

$$\tilde{r}(t) = \gamma_1 \hat{r}(t) + \epsilon.$$

In a linear regression model, removing the intercept term forces the model to go through the origin (point $(0{,}0)$). In doing so, the assumption is made that the dependent variable, in this instance the government-published estimates, is solely determined by the explanatory variables, the ONS-based estimates, and random noise. Thus, should these explanatory variables be 0, this suggests that the dependent variable is determined solely by noise. In this instance, we know that this is an incorrect specification of the relationship between our variables, as the government-published estimates were derived from an ensemble of models. However, we can use the no-intercept model to explore further the relationship between the ONS-based and government-published estimates. If $\widehat{\beta_1}\ or\ \widehat{\gamma_1}$ tend to 1, without the intercept, this implies that the ONS-based estimates are unbiased predictors of the government-published estimates. If $\widehat{\beta_1}\ or\ \widehat{\gamma_1}$ tend to 0, this implies that the government-published estimates are not predicted by our ONS-based estimates.

Supplementary Figure 11 presents the resulting $\widehat{\beta_1}$ and $\widehat{\gamma_1}$ for these models under the same lags (plus and minus 20 days) applied to the government-published estimates as in the main text. For $R(t)$, we see that the two time series have a strong relationship regardless of lag, with $\widehat{\beta_1}$ sitting consistently around 1. This is in contrast to $r(t)$, for which $\widehat{\gamma_1}$ approaches 1 at approximately the same lag shown to maximise $R^2$ in the models with intercepts in the main text.

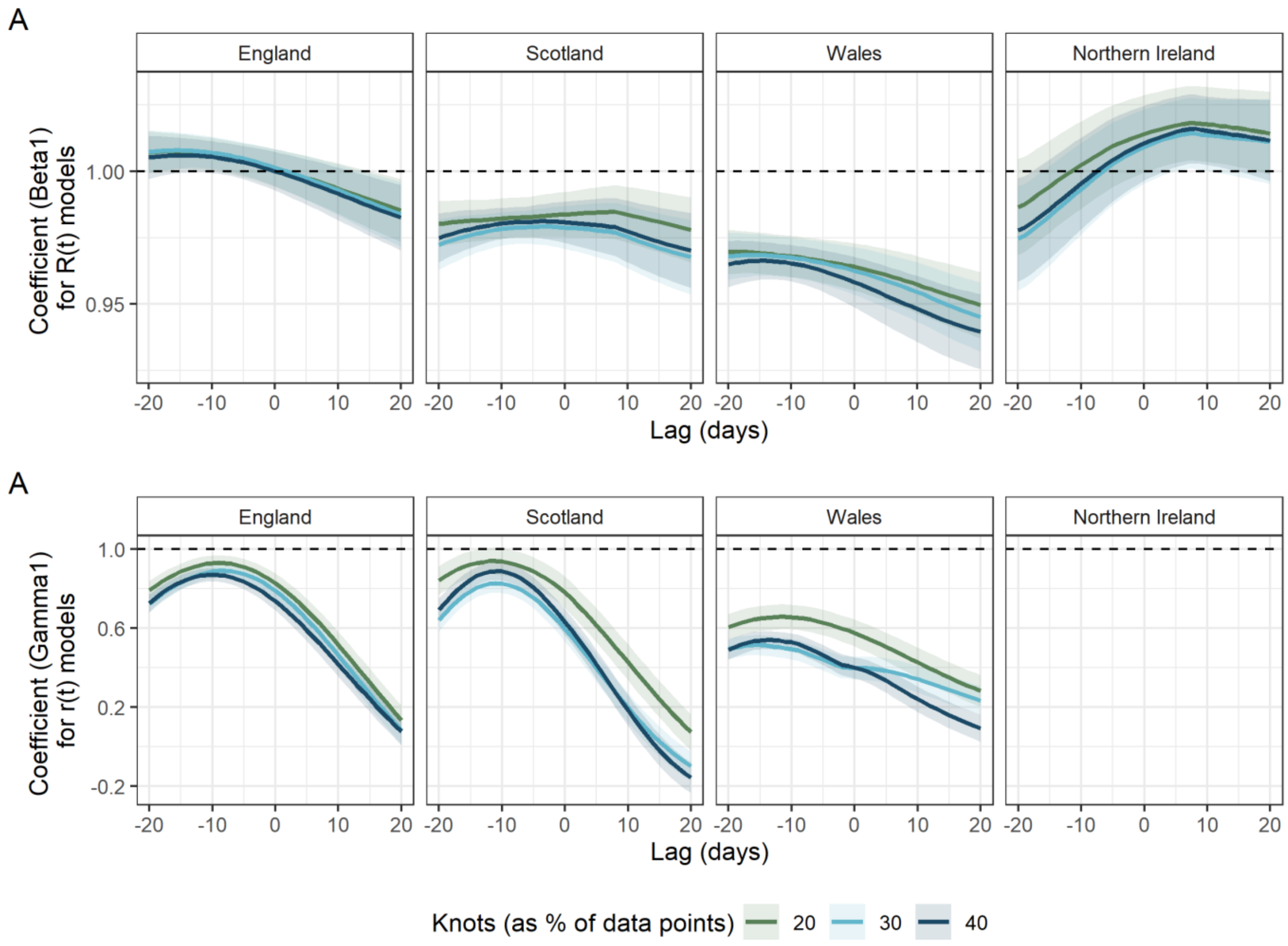

*Supplementary Figure 11: Coefficients (ordinary least squares (OLS) estimate (solid lines) with 95% confidence intervals (shaded areas) for $\widehat{\beta_1}$ and $\widehat{\gamma_1}$) from linear regression models regressing ONS-based estimates on lagged government-published estimates without the inclusion of an intercept, for England, Scotland, Wales and Northern Ireland under different levels of knots, given as a percentage of the total data points. Dashed line indicates coefficient value of 1. (A) Output for $R(t)$. (B) Output for $r(t)$. Northern Ireland is omitted due to the lack of government-published estimates for which to compare to.*

### References

1. Coronavirus (COVID-19) Infection Survey, UK: 6 January 2023. *Office for National Statistics* https://www.ons.gov.uk/peoplepopulationandcommunity/healthandsocialcare/conditionsanddiseases/bulletins/coronaviruscovid19infectionsurveypilot/6january2023 (2023).
